# A proteomic polygenic score to identify IL-18 driven inflammatory bowel disease

**DOI:** 10.64898/2026.05.18.26353508

**Authors:** Michael C Turchin, Narayanan Raghupathy, Cara L. Carty, Melody Morris, Joseph C Maranville, Emily R. Holzinger

## Abstract

High levels of IL-18 have been causally implicated in IBD risk and may represent a unique mechanism driving IBD yet to be therapeutically targeted. To identify individuals predisposed to increased levels of IL-18, we implemented a polygenic approach to predict IL-18 plasma protein levels. Using a dataset with over 50,000 individuals with both genetic and plasma protein levels from Olink, we developed a 27 SNP polygenic score that predicts IL-18 levels and IBD risk. Further, we identified a threshold to classify patients as ‘IL-18 High’ using a data-driven approach that optimized prediction of both IL-18 and IBD risk. We show that ∼30% of the overall IBD patient population is ‘IL-18 High’, meaning a genetic predisposition towards higher protein levels. The IL-18 PGS and corresponding threshold have the potential to identify IBD patients with IL-18-driven IBD that may respond more effectively to a therapy targeting this mechanism.

## Introduction

IBD is a heterogenous disease with evidence for multiple disease driving mechanisms^1–3^. One such mechanism may be increased IL-18 signaling, as high levels of IL-18 have been connected to the etiology of IBD through multiple sources of evidence^4^. First, circulating and intestinal IL-18 levels are higher in IBD compared to non-IBD patients^5–7^. Second, multiple IBD GWAS have identified a locus containing both IL-18Ra and IL-18Rb as significantly associated with IBD^8,9^. Third, Mendelian Randomization (MR) approaches have shown an association between increased IL-18 plasma protein and increased IBD risk, implicating a causal link between IL-18 and IBD risk^10^. And fourth, rare mutations that cause monogenic inflammatory disorders including very-early onset-IBD (VEO-IBD) present with increased IL-18^11–14^.

Specifically, gain-of-function mutations in NLRC4, an IL-18-inducing inflammasome component, lead to an autoinflammatory disorder that presents with enterocolitis and constitutively high IL-18 levels^11,12^. Further, loss-of-function mutations in XIAP, an antiapoptotic regulator of multiple immune pathways including IL-18, also lead to an autoinflammatory disorder whose presentations include IBD and high IL-18 levels^13,14^. Importantly, in both disorders, patients were only successfully treated after administration of recombinant human IL-18BP, a protein that binds and sequesters IL-18, to lower IL-18 levels^11,12,14^. All together, these pieces of evidence implicate IL-18 signaling as a driver of IBD risk.

From this evidence, we hypothesize that IBD patients with high IL-18 levels throughout their lifetime may benefit from IL-18 pathway inhibitors. However, identifying patients with sustained elevated levels of IL-18 is not straightforward. Assays measuring IL-18 levels alone only provide information about a single time point, and it is known that cytokines often vary due to a variety of biological and environmental factors^15–18^. Thus, using measurements of the cytokine level is insufficient for estimating lifetime exposure to high circulating IL-18. In this work, we propose a polygenic score (PGS) to predict IL-18 to address these difficulties.

PGS are tools that predict an outcome of interest by combining multiple SNPs^19–21^, resulting in a single value, or score, for any given individual that represents overall genetic risk due to common variants. These approaches are being taken beyond disease risk prediction and into proteomic prediction to capture causal mechanisms for disease, such as TIMP2 in Alzheimer’s^22^ and TLR1/IL27 in asthma^23^. With TIMP2, a proteomics-based PGS predictor of TIMP2 levels was associated with improved cognitive performance in individuals at risk for Alzheimer’s disease^22^; however, no association was observed between cognitive performance and circulating TIMP2 protein levels. This suggests lifelong exposure to higher TIMP2 may be driving the effect, a disease mechanism that can only be captured by a proteomic PGS and not by direct measurements of protein levels alone.

Furthermore, PGS may be a powerful tool for identifying the most appropriate patient populations for a specific therapy^24–26^. By identifying patient populations that would respond best to a given therapy, PGS could lower costs and increase success rates of clinical trials. For example in one study, retrospective analyses of two CVD-related trials found that individuals that were high for a PGS based on CAD risk and endothelial tissue function had a greater response to lipid-lowering therapies^27^. However, there are very few prospective clinical study designs that have incorporated a PGS for patient selection^28^. Therefore, understanding how disease mechanism-informed PGS relate to patient outcomes is crucial for effective implementation in clinical trials.

To understand the utility of PGS in targeting mechanistically-defined patient populations, we constructed and evaluated an IL-18 proteomics PGS to identify an IBD patient population genetically pre-disposed to higher IL-18 levels throughout their lifetime. We used the UK Biobank Pharma Proteomics Project (UKB-PPP) dataset^29^ to construct our PGS and validated it in an independent dataset of IBD patients. We then characterized the relationship between the IL-18 PGS and IBD-relevant endpoints in the full UKB^30^, beyond the participants in UKB-PPP, and IBD Plexus SPARC^31^ datasets. We used IL-18 levels and IBD risk to calibrate an optimal cutoff to determine PGS-high patients. We also implemented an approach that allows for PGS calculation on an individual level that is transferable across ancestries, which is crucial for clinical trial implementation.

## Results

### IL-18 GWAS, Conditional Mapping, and Mendelian Randomization

To create an IL-18 proteomics PGS, we first constructed a genetic map of all IL-18 associations using the UKB-PPP Olink dataset. We used the full UKB-PPP cohort, which included >50k individuals of predominantly European ancestry (>90% European ancestry) and ∼14M variants post-QC^32^. Additionally, to create the most accurate genetic map of IL-18 protein levels, we adjusted IL-18 levels by IL-18BP protein levels. IL-18BP binds and sequesters circulating IL-18^33^; Olink data, however, returns “total IL-18” levels, not “free IL-18” that is unbound and signaling. Therefore in lieu of a direct measurement of free IL-18, we hypothesized that accounting for IL-18BP levels may at least partially account for this discrepancy. Lastly, to derive a complete genetic map of IL-18 protein levels, we conducted full conditional mapping of our initial IL-18 GWAS (see Materials and Methods for details). In total, we found 27 significant associations between SNPs and IL-18BP-adjusted IL-18 NPX levels (Figure 1 and Table S1; 18 lead SNPs and 9 conditional SNPs at an association *p*-value < 3.55E-09).

**Figure 1.**
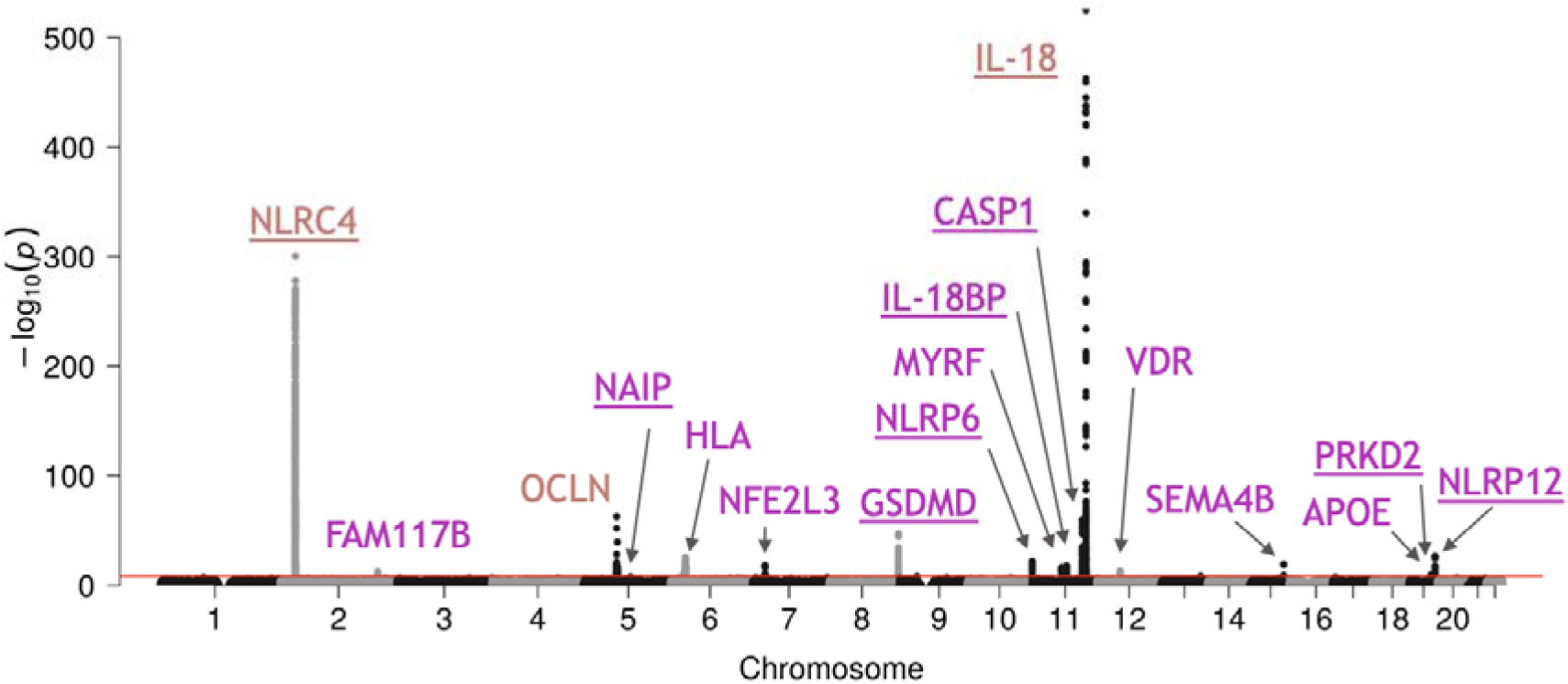
IL-18 Genetic Map Adjusted For IL-18BP. A Manhattan plot showing the results of our Olink IL-18 GWAS using a model that is adjusted for Olink IL-18BP levels. SNPs are ordered by chromosome and basepair position along the *x*-axis. The negative log10 of the association *p*-values are shown along the *y*-axis. Beige text indicates the previous three loci identified by Mokry et al. 2019^10^. Purple text indicates new loci identified beyond those three. Underline indicates gene has a direct connection to either the IL-18 pathway or inflammasome biology.

Across these 27 SNP associations, components of the IL-18 pathway and inflammasome machinery were represented. These include IL-18, IL-18BP, NLRC4, NAIP, CASP1, GSDMD, NLRP6, NLRP12, and PRKD2. IL-18BP was previously detailed. NLRC4, a member of the NLR-family of proteins and previously mentioned, defines one of the main inflammasomes that activates IL-18^4,34^. Similarly, CASP1 is also a component of the NLRC4-inflammasome and cleaves the pro-IL18 form of IL-18 to activate it^4,35^. Note that the CASP1 locus also includes other IL-18/inflammasome relevant genes, including CARD16/CARD17/CARD18 (putative inhibitors of CASP1^36^). NAIP is a pathogen sensing protein in the NLRC4-inflammasome as well^37,38^. GSDMD, part of the gasdermin family of proteins, is required for pyroptosis and is cleaved by CASP1 via induction from NLRC4- and other inflammasomes^39,40^. NLRP6 and NLRP12 are components of other inflammasomes that have evidence of promoting IL-18^41–44^. And lastly, PRKD2 is a member of a protein kinase family that has been found to regulate NLRP3^45,46^.

Despite many of these loci being trans-pQTLs, we hypothesized that with such strong connections to IL-18 and inflammasome biology, these additional loci may strengthen the evidence that high IL-18 levels are causally linked to IBD risk. To test this hypothesis, we conducted an MR analysis using our 27 lead SNPs similar to the MR analysis conducted by Mokry et al. 2019^10^. We used GSMR^47^ to do a multi-SNP MR analysis of IL-18 levels (exposure) and IBD risk (outcome), and indeed found a stronger associations between higher levels of IL-18 and IBD risk than what was previously seen^10^ (GSMR *p*-value 2.08E-06 vs. 1.07E-4, respectively; Figure 2 and Table 1). Additionally, in further support of our approach to adjust by IL-18BP levels, we see that a GSMR analysis using unadjusted IL-18 levels against IBD actually produced a less significant result (GSMR *p*-value 3.30E-05 vs. 2.08E-06, respectively; Table 1). Note that for the remainder of this manuscript, ‘IL-18 levels’ continues to refer to ‘IL-18BP-adjusted IL-18 levels’ unless otherwise indicated.

**Figure 2.**
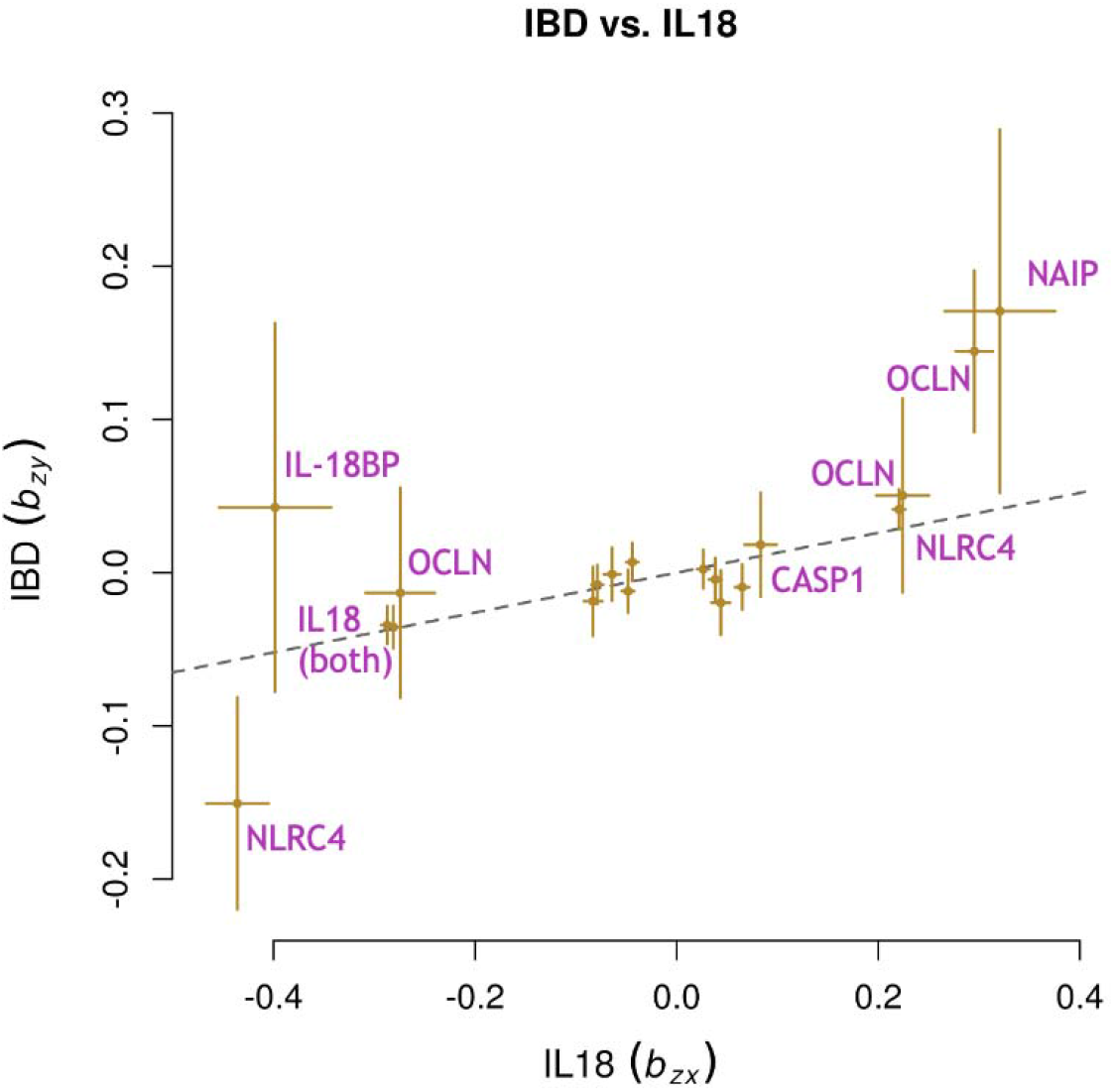
SNP associations from IL-18BP-adjusted model with Olink IL-18 and IBD underlying GSMR results. A scatter plot showing associations of the lead SNPs from the IL-18 GWAS with Olink IL-18 (adjusted for IL-18BP) and IBD risk. Associations between these SNPs and Olink IL-18 (adjusted) are shown on the *x*-axis and associations between these SNPs and IBD risk are shown on the *y*-axis. Olink IL-18 (adjusted) association information comes from the UKB-PPP IL-18 GWAS (adjusted) conducted internally. IBD risk association information is based on summary statistics from de Lange et al. 2017^8^. Bars on each SNP going along the *x*-axis and *y*-axis represent standard errors for each association. Purple text indicates loci represented by SNPs with stronger associations.

**Table 1.**
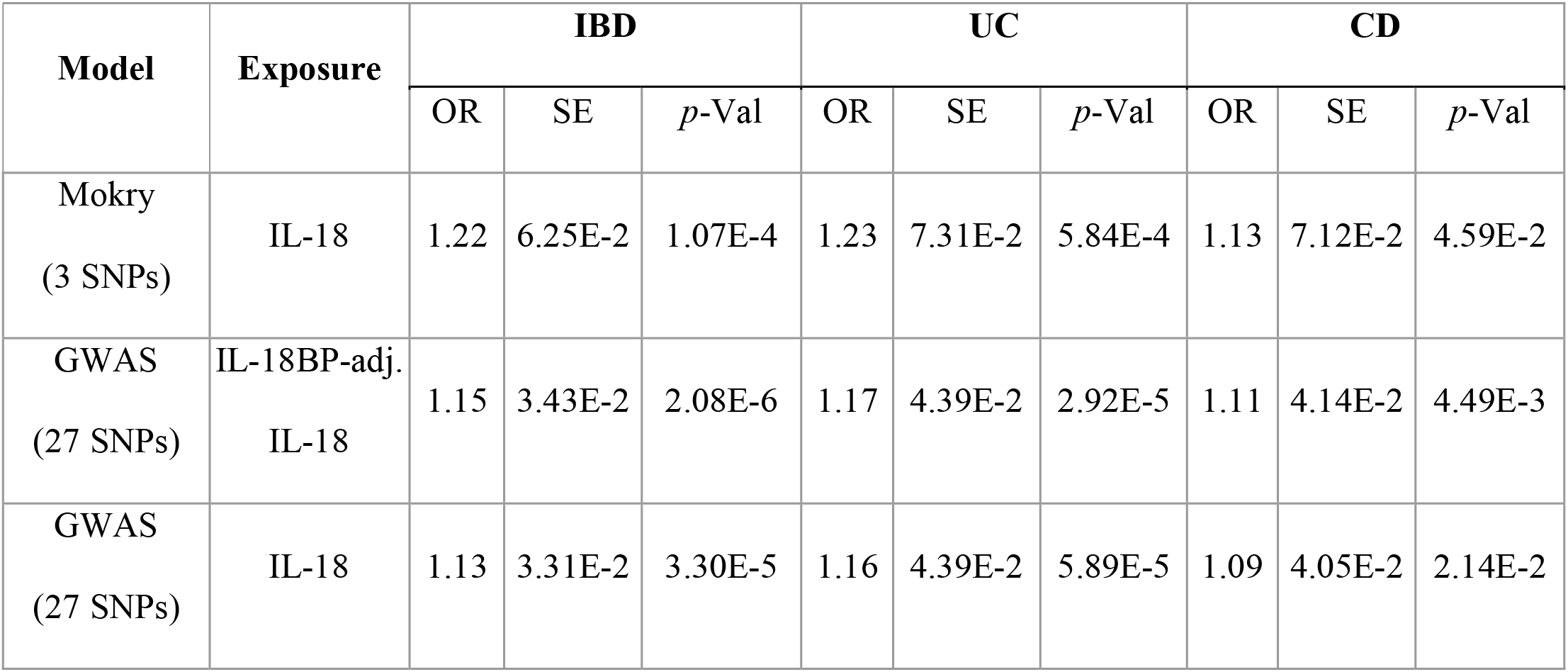
Investigating the causal link between high IL-18 and IBD risk using GSMR and different models. The table shows results from running GSMR, a multi-SNP MR method, to look at impact of Olink IL-18 levels on IBD risk. Results from three different models are being shown, either the 3 SNPs and data used by Mokry et al. 2019^10^, the 27 lead SNPs identified from the IL-18 GWAS and conditional analyses conducted internally, and the 27 lead SNPs identified from the IL-18 GWAS adjusted for IL-18BP and conditional analyses conducted internally. IL-18 association information comes from either the IL-18 GWAS or the IL-18 adjusted for IL-18BP GWAS conducted internally. IBD, UC, and CD risk association information comes from the IBD, UC, and, CD GWAS of de Lange et al. 2017^8^. Results are from using GSMR HEIDI outlier analysis. Note that due to SNP overlap between the UKB-PPP GWAS and the de Lange GWAS data, an initial set of 25 of 27 SNPs and 26 of 27 SNPs were used for the IL-18 and IL-18-adjusted for IL-18BP PGS analyses, respectively.

### IL-18 PGS Construction and Validation

Having further strengthened the evidence for a causal link between high IL-18 and IBD risk, we constructed a polygenic predictor for IL-18 protein levels. Using this IL-18 GWAS as our discovery dataset, we compared multiple approaches to construct an IL-18 PGS, including PRSice2^48^, full conditional mapping (FCM), LDpred2^49^, Lassosum^50^, SBayesR^51^, and PRS-CS^52^. To validate and compare the different PGS approaches, we used IBD Plexus SPARC, an independent dataset of 731 IBD patients that have both genetic and Olink information. We ran each PGS model in IBD Plexus SPARC. Across all six approaches, the constructed PGS significantly predicted IL-18 levels (Table 2; we also included the Mokry 3 SNP model for comparison). However, the most significant association came from the FCM PGS, where we used each lead SNP from every primary and conditional association (see Figure S1 for SPARC scatterplot of FCM PGS against IL-18 NPX values). We therefore focused on the FCM model as our choice for the PGS.

**Table 2.** Validation of IL-18 PGS models constructed through multiple standard approaches using IL-18BP-adjusted IL-18. The table shows results from validating various IL-18 PGS models by testing how these PGS predict Olink IL-18 (adjusted for IL-18BP) in the IBD Plexus SPARC dataset. Models were constructed using the different approaches detailed in the first column with the UKB-PPP IL-18 GWAS results as the initial discovery dataset. See Materials and Methods for details on how each method was implemented. Models were validated by testing their ability to predict Olink IL-18 NPX (adjusted for IL-18BP) in an independent dataset, the IBD Plexus SPARC dataset. IL-18 NPX vs. PGS beta and *p*-Value refer to the beta and *p*-value associated with the PGS coefficient in the full regression model: IL-18 NPX ∼ PGS + Age + Sex + Top 10 PCs; note that the beta is from using a standardized version of each PGS to make results comparable across methods. PGS-Only R2 refers to the total IL-18 NPX variance explained by the PGS model in a model that only contains the PGS as a predictor: IL-18 NPX ∼ PGS.

| IL-18 PGS Model | SNPs | Beta | *p*-Value | PGS-Only R2 |
| --- | --- | --- | --- | --- |
| Mokry 3 SNPs | 3 | 0.220 | 1.98E-10 | 0.049 |
| PRSice2 (C+T) | 181 | 0.221 | 3.93E-10 | 0.056 |
| FCM | 27 | 0.266 | 9.71E-15 | 0.080 |
| LDpred2 | 1.16E+06 | 0.242 | 1.97E-12 | 0.068 |
| Lassosum | 1.16E+06 | 0.250 | 1.13E-12 | 0.067 |
| SBayesR | 9.28E+05 | 0.226 | 1.73E-07 | 0.034 |
| PRS-CS | 8.97E+05 | 0.238 | 4.52E-12 | 0.066 |

### IL-18 PGS Characterization

Next, we tested for an association to IBD risk. First, we used an independent set of UKB individuals from the UKB-PPP dataset to evaluate the association between the IL-18 PGS and IBD, UC, and CD. Using ICD9/10 codes and self-report to assign case/control status following other publications^53,54^, we identified 3,987/360,107 IBD cases and controls, 2,707/361,387 UC cases and controls, and 1,463/362,631 CD cases. We then analyzed whether the IL-18 PGS significantly predicted IBD, UC, or CD. We found that the IL-18 PGS significantly predicts IBD and UC but not CD (Table 3).

**Table 3.** Association between the IL-18 PGS and IBD risk in the UKB. The table shows results from testing the association between the IL-18 PGS and IBD, UC, and CD risk in the UKB. IBD, UC, and CD status was determined using ICD9/10 codes and self-report. Case and control numbers are shown in the first column under each disease condition, and the OR and the *p*-Value from each association analysis are shown in the second and third columns.

| Dataset | IBD |  |  | UC |  |  | CD |  |  |
| --- | --- | --- | --- | --- | --- | --- | --- | --- | --- |
|  | Cs/Cntrl | OR | *p*-Value | Cs/Cntrl | OR | *p*-Value | Cs/Cntrl | OR | *p*-Value |
| UKB | 3982/<br>360107 | 1.08 | 2.31E-06 | 2707/<br>361387 | 1.1 | 1.23E-06 | 1463/<br>362631 | 1.03 | 2.33E-01 |

We next analyzed whether partitioning the IL-18 PGS into bins would reveal an enrichment of IBD, UC, or CD cases among the tail-end of the PGS spectrum. Breaking the IL-18 PGS into deciles, we do indeed see more significant odd ratios among the tail-ends of the PGS for both IBD and UC (Figure 3 and Figure S2). Though similar to before, we also see a weaker signal in CD. Taken together, these results suggest that our IL-18 PGS is identifying a subset of the IBD patient population that is related to high IL-18 levels.

**Figure 3.**
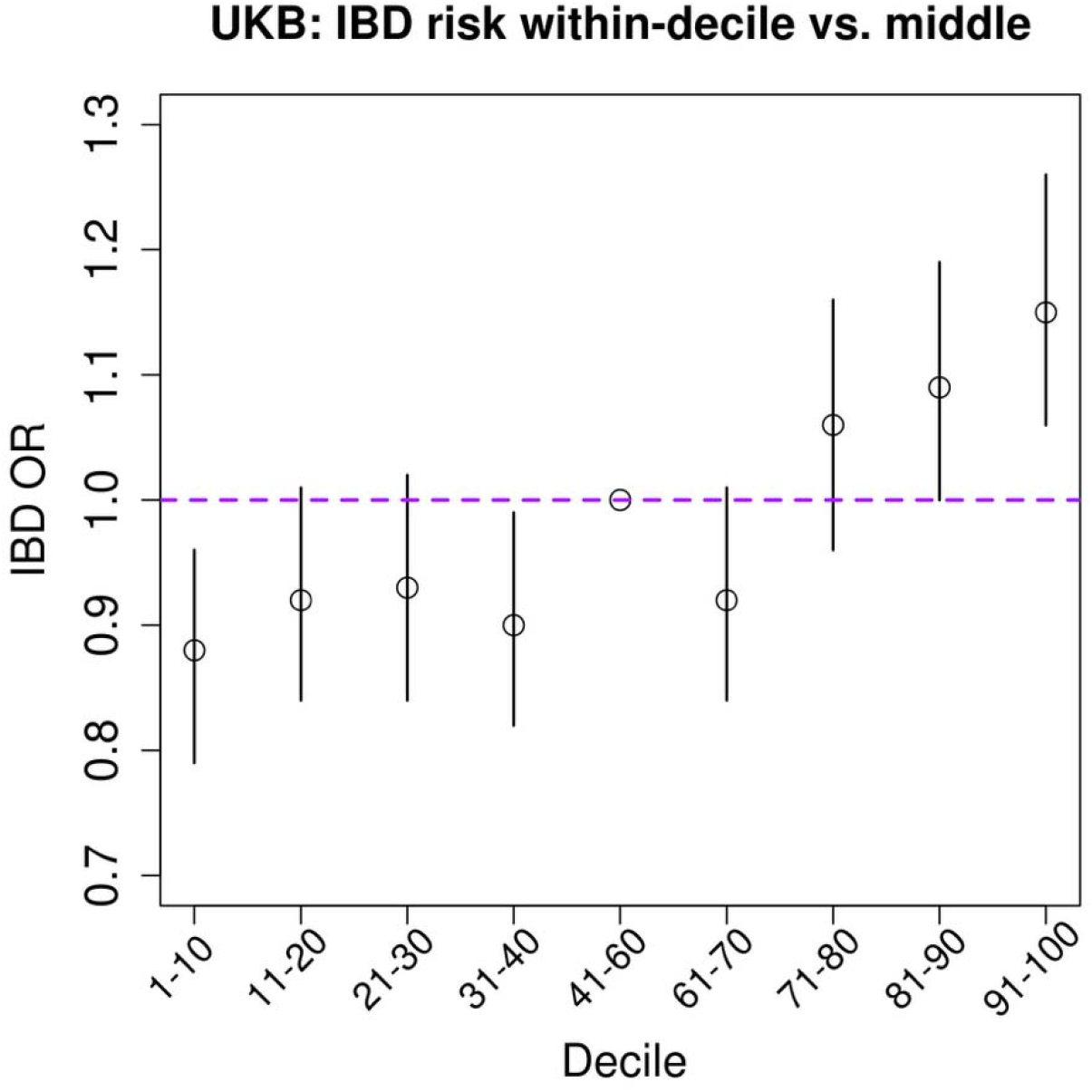
Prediction of IBD using IL-18 PGS Deciles. A decile plot showing the association between different deciles of the IL-18 PGS and IBD risk when compared to the middle deciles of the dataset. The different deciles of the IL-18 PGS are shown across the *x*-axis. The odds ratio of the association between each decile and IBD risk when the middle deciles of 41-60% are treated as the PGS control population are shown on the *y*-axis. Analysis was conducted in the non-UKB-PPP portion of the UKB and IBD case status was determined using ICD9/10 codes and self-report (see materials and methods). Error bars represent the 95% confidence intervals. The full model tested was as follows: IBD status ∼ IL-18 PGS decile status + Age + Sex + Top 10 PCs.

We further leveraged the IBD Plexus SPARC dataset to look at whether the IL-18 PGS was associated with IL-18 gene expression levels or clinical characteristics. For 327 UC and CD genotyped individuals from the SPARC dataset that also had gene expression data from colon biopsies from within 20cm of the anal verge, we found that the PGS significantly predicts IL-18 gene expression (beta: 0.237, *p*-value: 1.28E-05; Figure S3 and Table S2). This suggests individuals with a genetic predisposition to high IL-18 protein levels may also be experiencing high IL-18 gene expression levels in disease relevant tissue.

Next, we estimated associations between the PGS and disease activity in UC cases using Mayo-6 scores and in CD cases using short-CD activity index (sCDAI) score categories. In models adjusted for age, sex, genetic ancestry principal components and years since diagnosis at time of the disease activity assessment, we did not find significant associations between the PGS and binary outcomes (Remission vs. Mod/Severe or Remission/Mild vs. Mod/Severe) in either UC or CD cases (Table 4). These null findings suggest that standard IBD disease activity scores may not reflect the disease activity most relevant to an IL-18 driven IBD patient population.

**Table 4:** IL-18 PGS Associations with Disease Activity as Measured by 6pt Mayo Score for UC and sCDAI for CD. The table shows results from comparing the IL-18 PGS between different levels of IBD disease activity as measured by the 6pt Mayo (UC) or short Crohn’s disease activity index (sCDAI) score classifications. Analyses were conducted in the IBD Plexus SPARC dataset. Patients were grouped as either Moderate or Severe combined, Remission and Mild combined, or only Remission. The binomial model used was: Disease Activity Group ∼ IL-18 PGS + Age + Sex + Top 10 PCs + Years since diagnosis at time of disease activity measurement. A model treating Disease Activity as an ordinal outcome produced similar results (data not shown).

| Case Group | IL-18 PGS and Disease Severity Categories | *p*-Value |
| --- | --- | --- |
| CD | Remission (n=542) vs. Mod/Sev (n=503) | 0.733 |
|  | Remission/Mild (n=899) vs. Mod/Sev (n=503) | 0.265 |
| UC | Remission (n=301) vs. Mod/Sev (n=165) | 0.395 |
|  | Remission/Mild (n=503) vs. Mod/Sev (n=165) | 0.783 |

We also tested the relationship between the IL-18 PGS and TNF-a treatment failure using an algorithmic approach to derive treatment failure in CD and UC cases. While no significant association between the PGS and TNF-a treatment failure was observed in CD (*p-*value: 0.762) or UC (*p*-value: 0.436) cases in covariate adjusted models, we acknowledge limited power to detect modest effects in these analyses. TNF-a treatment failure status was limited to 181 UC cases (Yes/No N: 49/132) and 571 CD cases (Yes/No N: 201/370), which is a smaller subset of the overall IBD Plexus SPARC cohort.

### IBD Risk-Informed IL-18 PGS Model

Having observed the connection between the IL-18 PGS and IBD risk, we wanted to explore a different PGS model that takes IBD risk direction into account for SNP selection. We created a PGS model based on our previous MR analysis that only included SNPs where the IL-18 increasing allele was also associated with increased IBD risk (based on de Lange et al. 2017^8^). Doing so produced an 18 SNP model. Comparing this ‘IBD risk’ version of our PGS model with the previously generated IL-18 PGS, we find similar prediction of IBD risk (4.00E-06 vs 2.31E-06 *p*-values for IBD prediction; Table S3). In terms of IL-18 prediction however, the IL-18 PGS model outperforms the ‘IBD risk’ PGS model, likely due to inclusion of more IL-18-associateed SNPs (9.19E-13 vs 9.71E-15 *p*-values for IL-18 prediction; Table S3). Given the similar performances in IBD risk prediction but differences in IL-18 prediction, we continued with the IL-18 PGS model. However, this is a further example of how one might generate a PGS informed by disease mechanism and is worth continued exploration.

### IL-18 PGS Threshold Calibration

With an IL-18 PGS that predicts both IL-18 protein levels and IBD risk, we used a data-driven approach to calibrate a threshold to categorically define IL-18 high individuals. To do this, we used a grid search approach to identify an optimal PGS cutoff for both high IL-18 individuals as well as IBD risk. Additionally, we wanted this PGS cutoff to be transferable across ancestries and datasets; to accomplish this, similar to previous approaches^55,56^, we used PCA projection and the 1000 Genomes as an external reference dataset to correct PGS by ancestry.

First, we used the 1000 Genomes as a reference dataset to generate a PC space using ∼450k lightly pruned SNPs. We next projected individuals from either the UKB or IBD Plexus SPARC into this space such that we could calculate PCs on every individual separately (as would be the case in a clinical trial). Lastly, each individual in the UKB or SPARC dataset had their PGS standardized using means and variances predicted from trained linear models using these projected PCs (see methods for further details). Doing this helps make the expected percent of individuals designated as PGS-high given a cutoff more consistent across ancestry groups and allows single individuals to have their PGS values corrected properly without needing a full genetic dataset on hand.

Second, once PGS models for each individual were standardized, the optimal PGS threshold for high IL-18 prediction was determined using the IBD Plexus SPARC dataset. We created binary high/low IL-18 PGS predictors using a range of cutoffs from 0.01 to 0.99 percent and tested how well these new binary variables predicted IL-18 Olink levels. Additionally, we simulated the presence of a real IL-18 high population by splitting individuals into IL-18 high/low categories using the same cutoff range of 0.01 to 0.99 percent. This created a 99 by 99 grid-search where for every IL-18 protein level cutoff we iterated over each IL-18 PGS cutoff in the ranges described above. Looking across all 9,801 combinations of cutoffs, we found that an IL-18 PGS threshold of 0.52 percent produced the most significant prediction of IL-18 high/low Olink levels (Figure S4A).

Third, to identify the optimal PGS threshold for IBD risk, we returned to using the previously created UKB subset that was independent of UKB-PPP individuals. We took the same approach as with the IL-18 prediction grid-search, creating binary high/low IL-18 PGS predictors using cutoffs from 0.01 to 0.99 percent and seeing how well these new binary variables predicted IBD risk. Once again, looking across the 99 cutoffs used, we found that a threshold of 0.80 produced the most significant association with IBD risk (Figure S4B).

Lastly, to combine these two thresholds into a single IL-18 PGS cutoff, we took the average of both. This produced a final cutoff of 0.66 (Figure 4), which would lead to 34% of the population being designated as IL-18 PGS high. We note that this final cutoff ends up aligning with the top 30% of IL-18 PGS deciles that are associated with (or trending towards) IBD risk in Figure 3.

**Figure 4.**
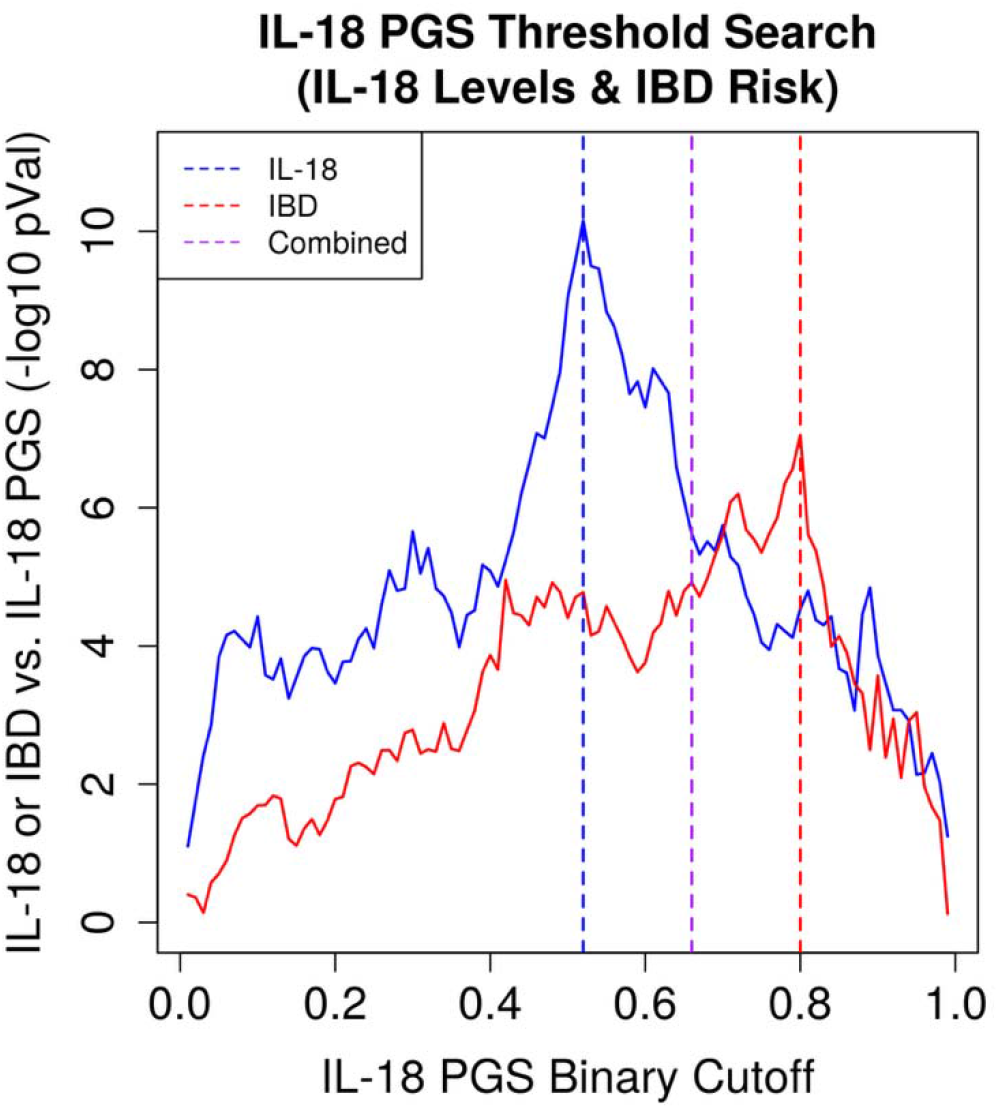
IL-18 PGS High Threshold Grid-Search Results. A line graph showing the association between binarized versions of the IL-18 PGS and either Olink IL-18 levels or IBD risk across a range of IL-18 PGS high/low cutoffs. The range of cutoffs used to convert the IL-18 PGS into high/low bins, incremented by 0.01, is shown on the *x*-axis. The negative log10 *p*-values of the association between the binarized IL-18 PGS predictors and either Olink IL-18 levels or IBD risk is shown on the *y*-axis. Olink IL-18 information comes from IBD Plexus SPARC and IBD risk information comes from the non-UKB-PPP subset of the UKB dataset. Blue dotted line represents a cutoff at 0.52 based only on Olink IL-18. Red dotted line represents a cutoff at 0.80 based only on IBD risk. Purple dotted line represents a combined cutoff at 0.66 based on both Olink IL-18 and IBD risk.

Given this final cutoff value, we also confirmed that our PCA projection approach was indeed making this threshold more transferable across human ancestries. We compared the percentage of individuals that were called IL-18 high within each UKB ancestry subset using this 34% cutoff and our PGS correction approach. We also compared against other correction approaches, including not correcting the PGS at all, using directly calculated within-dataset PCs, using projected PCs based on a set of 128 ancestry informative markers, and using the genome-wide projected PCs without standardization. And indeed, we find that our approach leads to more consistency in the percent of individuals being called IL-18 high across human ancestries while using PC projection (Table 5 and Figure S5). Therefore, we identified an IL-18 PGS cutoff that would determine genetically-high IL-18 IBD patients across different human ancestries.

**Table 5:** IL-18 PGS High Cutoff Percentages Across Ancestries. The table shows the percent of individuals that are considered IL-18 PGS high across ancestry groups given the cutoff pertaining to 34% in each PGS model described. Each model corresponds to a different approach for correcting the IL-18 PGS. ‘No Adj’ refers to a model where the PGS is not corrected at all, ‘Age/Sex/10PCs’ refers to a model where age, sex, and top 10 PCs are regressed out from the IL-18 PGS directly within the UKB dataset, ‘Seldin128 (Direct)’ refers to a model where age, sex, and top 10 PCs are regressed out from the IL-18 PGS directly but the PCs are based on the 128 ancestry informative markers from Kosoy et al. 2009^64^, ‘Selding128 (Proj)’ refers to a model where age, sex, and top 10 PCs are regressed out but the top 10 PCs are based on projecting UKB individuals into a PC space generated using the 128 Seldin SNPs and 1000 Genomes, ‘450k SNPs (Proj)’ refers to a model where age, sex, and top 10 PCs are regressed out but the top 10 PCs are based on projecting UKB individuals into a PC space generated using 450k lightly pruned SNPs and 1000 Genomes, and ‘450k SNPs (Proj+)’ refers to a model that is similar to ‘450k SNPs (Proj)’ but the IL-18 PGS is standardized using predicted means and standard deviations from the trained linear regression models as described in Results and Materials and Methods. UKB ancestry groups are based on self-reported ancestry.

| **PGS Model** | **Expected**<br>**PGS+ %** | **African**<br>**(n=2447)** | **British**<br>**(n=318469)** | **Caribbean**<br>**(n=3365)** | **Chinese**<br>**(n=1291)** | **Indian**<br>**(n=4625)** | **Irish**<br>**(n=9538)** | **Pakistani**<br>**(n=1436)** |
| --- | --- | --- | --- | --- | --- | --- | --- | --- |
| No Adj | 34% | 52.3% | 33.1% | 48.1% | 59.6% | 45.4% | 33.9% | 49.0% |
| Age/Sex/10PCs | 34% | 39.4% | 34.0% | 36.8% | 29.1% | 33.3% | 33.6% | 36.4% |
| Seldin128 (Direct) | 34% | 40.0% | 33.7% | 36.4% | 35.9% | 39.7% | 34.7% | 42.8% |
| Seldin128 (Proj) | 34% | 45.3% | 33.4% | 41.5% | 53.8% | 43.1% | 34.3% | 46.9% |
| 450k SNPs (Proj) | 34% | 39.8% | 33.9% | 37.3% | 31.1% | 33.5% | 34.7% | 36.6% |
| 450k SNPs (Proj+) | 34% | 38.8% | 33.9% | 36.2% | 34.1% | 34.7% | 34.7% | 37.8% |

### IL-18 PGS Ancestry and SNP Sensitivity Analyses

Finally, using the datasets available to us, we aimed to evaluate how well this PGS predicts IL-18 levels in non-European populations. While the UKB-PPP dataset does have non-European ancestry individuals, it is predominantly European-ancestry, and the challenges with transferring single-ancestry-derived PGSs to other human ancestries is well established^57–59^. We therefore sought to evaluate how well this IL-18 PGS performs in non-European human ancestries. To accomplish this, we used both the UKB-PPP dataset as well as global allele frequency information from gnomAD^60^ and looked at both overall IL-18 PGS performance metrics and per-SNP performance metrics.

First, we created ancestry-specific subsets from the UKB-PPP dataset to get a sense of how the IL-18 PGS performs in ancestries beyond European individuals. We created three equally sized subsets (n=763) representing European, African, and Asian ancestries. Analyzing how well the IL-18 PGS predicts IL-18 levels in each of these three subsets, we find that the European and African ancestry subsets have similar performances and that the Asian ancestry subset has decreased performance (Figure 5). While all three subsets still significantly predict IL-18, these analyses are using the same individuals as the ones from the discovery IL-18 GWAS, meaning the results are biased towards overperforming (Figure 5).

**Figure 5.**
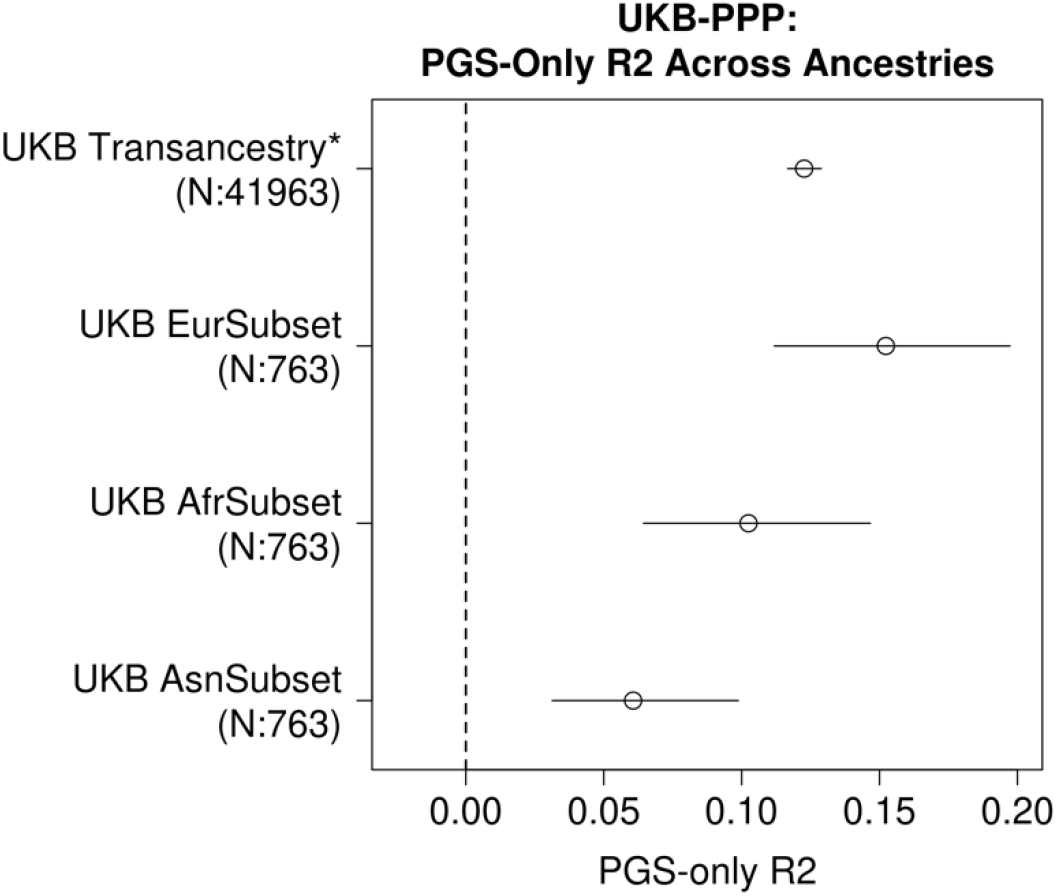
IL-18 PGS Performance Across Ancestry Subsets. The plot shows the predictive performance of the IL-18 PGS performance across different ancestry groupings in the UKB-PPP. The *x*-axis shows the IL-18 NPX (adjusted for IL-18BP) phenotypic variance explained by only the PGS. The *y*-axis shows the different ancestry groupings that were created: ‘UKB Transancestry’ refers to using all individuals from the original UKB-PPP IL-18 GWAS (adjusted for IL-18BP; related samples were also dropped), ‘UKB EurSubset’, ‘UKB AfrSubset’, and ‘UKB AsnSubset’ refer to three equally-sized, randomly sampled UKB-PPP subsets created to represent European, African, and Asian ancestries respectively (see materials and methods for more details). Sample sizes for each dataset used are also included on the *y*-axis labels. Data point 95% confidence intervals were determined with 10,000 bootstraps. The dotted vertical line represents a PGS-only r^2^ of 0.

To understand how the IL-18 PGS performs in each ancestry subset, as well as the importance of each SNP to the IL-18 PGS model, we looked at the impact of each SNP on the IL-18 PGS. We did this in two different ways. First, we looked at the performance of the IL-18 PGS in predicting IL-18 levels when taking a leave-one-out approach for each SNP in both the original full dataset as well as each ancestry subset. For almost every SNP, removal leads to a decrease in PGS-only R^2^ in the full dataset. Interestingly, one SNP actually leads to an increase in PGS-only R^2^ (rs1946518; Figure 6A). Looking at the ancestry subsets however, we see that this SNP is particularly important to the African and Asian subsets despite having worse performance in the European subset (Figure 6B). This suggests that despite the strong bias of the full datasets towards European ancestries, the IL-18 PGS model is still able to incorporate SNPs that are important to other non-European ancestries as well. This is further revealed when we take our second approach and look at global allele frequencies from gnomAD (Figure S6); we find that, while common across global ancestry groups, rs1946518 is at increased frequency in some African and Asian ancestry subsets, confirming its greater importance to multiple non-European ancestry groups. Overall, these results reveal that while it is likely the IL-18 PGS is still impacted by the European-ancestry bias of the UKB-PPP, the PGS is capturing relevant non-European ancestry information that should help improve its consistency across human ancestries.

**Figure 6.**
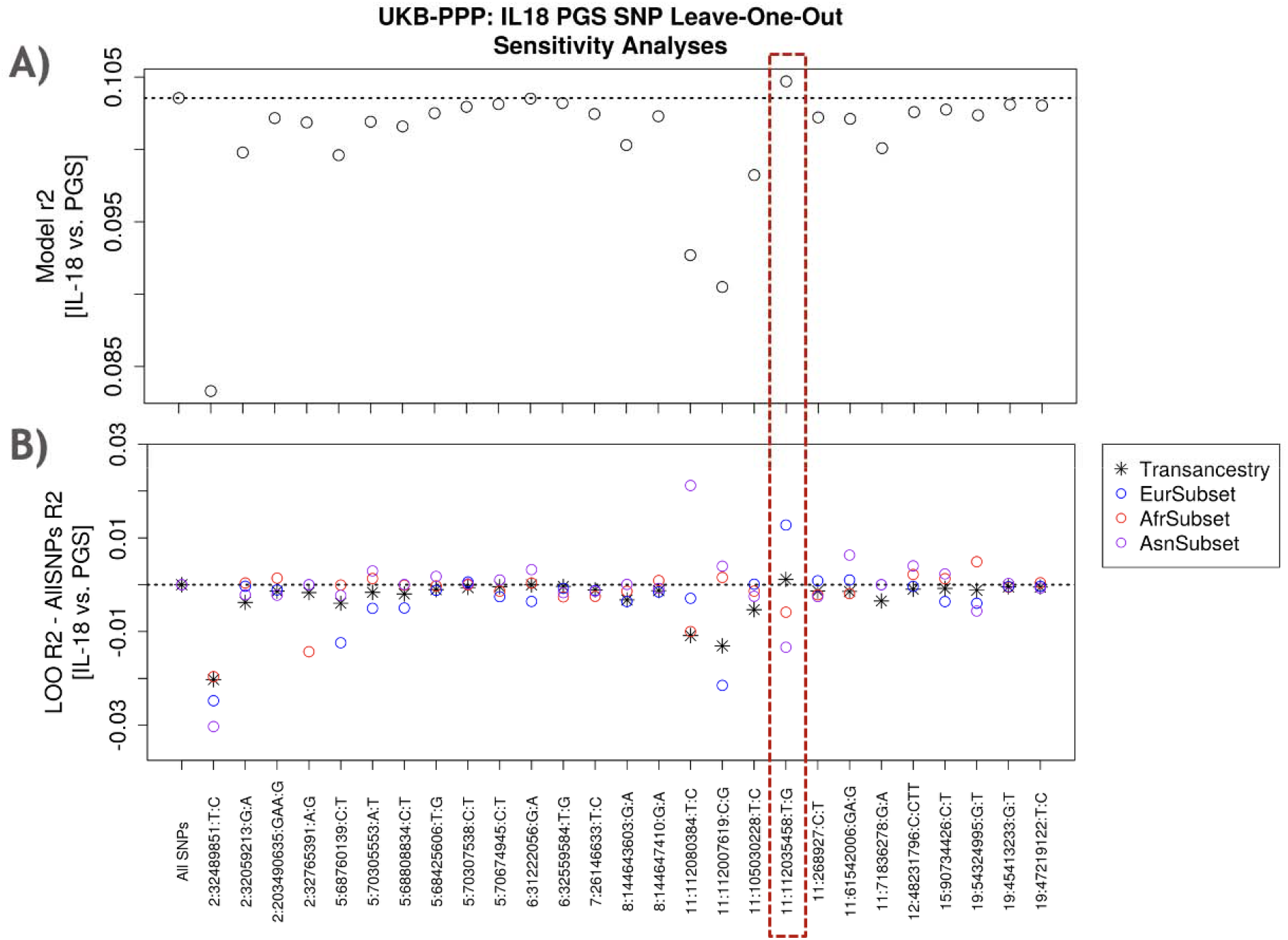
IL-18 PGS Single SNP Sensitivity Analyses Across Ancestry Groups. Both plots look at the sensitivity of IL-18 PGS predictive performance to the removal of each SNP in a leave-one-out manner from the full model. **A**) The first plot looks at the IL-18 NPX (adjusted for IL-18 BP) phenotype variance explained in the UKB-PPP dataset when each SNP from the IL-18 PGS is removed one at a time. The *x*-axis shows first the original full model (“All SNPs”) for comparison and then each SNP from the model that is removed one at a time. The *y*-axis shows the PGS-only phenotypic variance explained by each PGS model with the SNP indicated along the x-axis removed. The dotted horizontal line shows the r^2^ from the full model as a point of comparison. The dotted red rectangle highlights the SNP (rs1946518) where PGS-only r^2^ increases in the full UKB-PPP dataset when it is removed. **B**) The second plot looks at the change in IL-18 NPX (adjusted for IL-18 BP) phenotype variance explained when each SNP is removed from the PGS across different ancestry subgroups. The *x*-axis once again shows which SNP is being removed from the IL-18 PGS. The y-axis shows the difference between the r^2^ in each new leave-one-out (“LOO”) PGS model versus the original full (“AllSNPs”) within each ancestry grouping. Ancestry grouping definitions follow those described in Figure 5 (and Materials and Methods). The dotted horizontal lines shows a change in r^2^ of 0.

## Discussion

In this paper we detail the construction and evaluation of an IL-18 proteomics PGS to identify IBD patients that have systemically high IL-18 levels. We generate our IL-18 PGS by first running an IL-18 GWAS using the UKB-PPP dataset and validate our PGS by using the IBD Plexus SPARC cohort. We characterize the IL-18 PGS’s ability to significantly predict IBD using the UKB dataset. We also characterize the relationship between the IL-18 PGS and both clinical characteristics and IL-18 gene expression levels in the gut using SPARC. We then show how to create a transferable PGS cutoff for determining IL-18 high individuals using the 1000 Genomes as an external reference dataset and both IL-18 and IBD risk information. Lastly, we evaluate the performance of our IL-18 PGS across human ancestries in terms of phenotypic variance explained and look at how much each SNP contributes to this performance. Overall, these efforts are an essential addition to the growing body of work investigating how to implement PGS for therapeutic drug development.

There are some key aspects of our IL-18 PGS to highlight. First, by using all the lead and conditional lead SNPs from our IL-18 GWAS, we created an ‘IL-18 pathway’ PGS. In doing this we incorporated both cis-pQTLs and trans-pQTLs, similar to other recent proteomic PGS work^22,23^. Multiple trans-pQTLs had direct connections to IL-18 and inflammasome machinery and contributed to the overall variance explained by the PGS (Figure 6). Second, we created a model that aimed to adjust for the amount of circulating IL-18 that was bound and not signaling using levels of IL-18BP; while this led to an increase in performance in predicting IBD disease risk, thus supporting our choice to move forward with this model, it was still an approximation and should eventually be experimentally validated. Third, since we captured the IL-18 pathway in a disease agnostic way, it potentially creates the opportunity to use this PGS for other disease indications beyond IBD as well.

Additionally, we calibrated our PGS threshold so that it would be more consistent across human ancestries. As is well established within the PGS field, human ancestries need to be directly considered when constructing and implementing PGS^57–59^. It should be noted that to evaluate the performance of our PGS across human ancestries, we used the same dataset that our discovery GWAS was based on (UKB-PPP). While this could bias our estimates, there is still a lack of publicly available datasets that are both diverse and contain multi-omic information; finding another dataset that was as diverse as the UKB-PPP that had both genetic and proteomic information was infeasible at the time of this work. However, this will ideally not be the case as more diverse biobanks continue to grow in size and available datatypes.

Lastly, our efforts here add to the small but growing work of evaluating how to implement PGS in clinical trials. Our approach for identifying a PGS cutoff for IL-18 high individuals is innovative and incorporates both IL-18 and IBD risk information. And while we ideally would have conducted our grid search approach on the same set of individuals, we were once again limited by the availability of datasets. Additionally, by taking a grid search approach where we prespecified which cutoff values to evaluate, it is possible that there are some in-between thresholds we overlooked that would have further strengthened our results. And lastly, we combined our two threshold results by taking the average value; other approaches for determining this final cutoff value will be evaluated in future work. Overall, however, this work adds an important example of how to implement PGS in the context of clinical trials. There remains still a major need for prospective clinical trials that explicitly use PGS in their study design. More examples and data are needed to advance the field and accelerate adoption of PGS in clinical trials.

## Materials and Methods

### IL-18 UKB-PPP GWAS & Full Conditional Mapping

The UK BioBank Pharma Proteomics Project (UKB-PPP) Olink dataset was used to conduct our IL-18 proteomics GWAS. This dataset has previously been described^29^, but in short, the project ran the Olink Explore panel (3072 proteins) on 54,219 individuals from the UK BioBank. Proteomics data was quality controlled and processed as previously described and used as delivered. Genetic data was quality controlled following standard approaches for the UKB dataset. Imputed SNPs were dropped if their imputation info score was less than 0.8, minor allele frequency was less than 0.01%, missingness was > 5%, their Hardy-Weinberg test p-value was < 1e-50 in just the British ancestry individuals, and were not flagged as high-quality across ancestries by the Pan-UKBB Project^61^. Individuals were dropped if they were flagged as having any sex chromosome aneuploidy, outliers for excess of heterozygosity or missingness, or excess relatedness. This produced a final dataset of 50,782 individuals and >14M SNPs.

The IL-18 GWAS was conducted using regenie v2.2.4^32^. The GWAS model used was INT Adjusted IL-18 NPX ∼ Age + Sex + Age^2^ + Age*Sex + Age^2^*Sex + Batch + Assessment Center + Array + Inflammation_tbms + Sample Selection + Top 10 PCs, where INT refers to rank-based inverse normal transformation, Batch is the UKB-PPP proteomics batch run, Assessment Center is the UKB assessment center where participant consented, Array is the UKB genotyping array (BiLEVE or Axiom), Inflammation_tbms is the ‘time between measurement and sampling’ (a UKB-PPP created covariate specific to each Olink panel), and Sample Selection is the reason for sample inclusion in the UKB-PPP (randomized baseline, specific partner request, or COVID study). ‘Adjusted IL-18 NPX’ refers to IL-18 NPX levels that have been adjusted by IL-18BP NPX levels. And prior to being used for adjustment, IL-18BP NPX levels were corrected by the Cardiometabolic_tbms value as well. Annotation of GWAS results shown in Figure 1 were based on a combination of known connections to the IL-18 pathway or inflammasome biology in the literature, OpenTarget L2G scores^62^ from other published IL-18 proteomic studies, and the closest gene; the first two categories took priority when the information was available.

Full conditional mapping was conducted by manually rerunning regenie on each chromosome that had an initial lead SNP. Regenie was rerun by including each lead SNP as a covariate, and then each conditional lead SNP as an additional covariate afterwards, until no single SNP remained that had a genome-wide significant association (*p*-value < 3.55E-09). Each lead SNP and conditional lead SNP that was used as a covariate was then also dropped from the set of SNPs being tested for each conditional regenie run.

### IL-18 and IBD Mendelian Randomization

Mendelian randomization analyses were conducted using GSMR^47^. IL-18 GWAS data (without or with IL-18BP correction) came from our internally derived UKB-PPP IL-18 analysis detailed above, and IBD GWAS information came from the most well-powered, publicly available IBD GWAS at the time. All lead and conditional lead SNPs from our IL-18 GWAS analysis were initially included. SNPs that had ambiguous alleles (A/T or G/C) or IL-18 SNPs that did not overlap with the IBD GWAS were dropped and proxy SNPs were identified where possible (r2 > 0.4). This lead to a total of 26 SNPs from the original set of 27 SNPs being represented in these analyses, with 13 requiring proxy SNPs. HEIDI-outlier analysis was also used for these GSMR analyses, leading to a further 7 SNPs being removed and a final set of 19 SNPs analyzed with GSMR. To compare against the Mokry et al. 2019^10^ analysis, the same three SNPs from their publication and the same initial GWAS for IL-18 and IBD were used as input to GSMR.

### IL-18 PGS Construction and Validation

IL-18 PGS were constructed using multiple approaches, including PRSice2^48^, full conditional mapping (FCM), LDpred2^49^, Lassosum, SBayesR, and PRS-CS. In all instances, our internal IL-18 UKB-PPP GWAS was used as the discovery dataset. For PRSice, version 2.3.3 and the default parameters were used; a range of clumping r^2^ values were tested going from 0.01 to 0.9, and a model using a clumping r^2^ of 0.3 and a p-value cutoff of 3.55E-09 produced the strongest predictor. For FCM, a PGS model was constructed where all the lead variants and conditional lead variants were used from the full conditional mapping done during the IL-18 GWAS. For LDpred2, the R function snp_ldpred2_auto() from the bigsnpr package (version 1.12.16) was used following recommended parameters from the provided LDpred2 vignette. The IL-18 GWAS was intersected with the HapMap3+ variant set linked to by the vignette as well. For Lassosum, the R function snp_lassosum2() from the same aforementioned bigsnpr package was used following the recommended setup from the same aforementioned vignette. For SBayesR, version 2.05beta of the GCTB package and the default parameters were used. For PRS-CS, version v1.1.0 and the default parameters were used; phi values of 1, 1E-2, 1E-4, and 1E-6 were tested, with a model based on a phi of 1E-6 producing the strongest prediction.

To test and validate all the above models constructed in an independent IBD dataset, IBD Plexus SPARC was used^31^. The IBD Plexus SPARC cohort is a longitudinal study of adults with IBD from the Crohn’s and Colitis Foundation and includes data from both periodic study visits and electronic health records (EHR). SPARC genotype data from an Illumina Infinium Global Screening Array was first quality controlled by dropping SNPs with minor allele frequency < 1%, missingness > 1%, and Hardy-Weinberg test p-value < 1e-50. Samples were then also dropped if genotype missingness was > 1%, if their genetic sex did not match their self-reported sex, and if they were PCA outliers. PCA outliers were determined using the bigsnpr package and the approach detailed in section of 3.4 Privé et al. 2020^63^; a cutoff of > 0.8 for their described statistic was used to drop individuals. Samples were also dropped in a greedy manner if they were more than 2^nd^ degree relatives using PLINK v2.00a and the ‘--king-cutoff’ function. Post-QC genotype data was then imputed using a pipeline based on the University of Michigan imputation server and with the 1000 Genomes Phase 3 reference panel. Lastly, SNPs with imputation info scores < 0.3 were also dropped. This produced a final dataset of 2163 individuals and >28.5M SNPs, and 25 of 27 SNPs remaining from the IL-18 PGS model (no proxies included).

For SPARC Olink data from the Explore 3072 assay, samples were quality controlled following standard approaches as outlined by the R package OlinkAnalyze’s vignette (from Olink). Samples were dropped if they were PCA outliers, had an excess of QC warning flags across all panels, had an excess of QC warning flags across the Inflammation panel, and if they had a warning flag specifically for the IL-18 assay. This produced a quality controlled Olink dataset of 2332 samples. Of these 2332 samples and the 2163 individuals that had genetics information, 1048 overlapped. And for 252 individuals within this set of 1048 samples, they had multiple samples. In these instances, a random sample was chosen per individual so that each individual was represented by a single sample in an unbiased way. This produced a final SPARC genetics and Olink dataset of 731 samples and individuals.

To validate each of the above models in the processed SPARC data, the following linear regression model was used: INT IL-18 ∼ PGS Model + Age + Sex + Top 10 PCs. INT IL-18BP was also included as covariate when appropriate.

### IL-18 PGS Characterization

The IL-18 PGS model was tested for predicting IBD, UC, and CD, using the UKB dataset. Specifically, the non-UKB-PPP component of the UKB dataset was used. The dataset was further quality controlled using standard approaches for the UKB, including dropping individuals that are >= 2^nd^ degree relatives, individuals with excess relatedness, individuals with chromosomal aneuploidy, and individuals with excess heterozygosity; this produced a final dataset of 364,094 individuals. IBD, UC, and CD, status was determined using ICD9 and ICD10 codes and UKB self-report following similar approaches^53,54^; specifically, ICD9 codes 555 + 556, 556, and 555, ICD10 codes K51 + K50, K51, and K50, and UKB self-report codes 1461, 1463, and 1462, were used for IBD, UC, and CD, respectively. An individual was considered a case for IBD, UC, or CD if they had at least two designations for each individual disease across both the ICD9/10 codes and UKB self-reports. This resulted in the following IBD, UC, and CD case/control designations: 3,987/360,107, 2,707/361,387, and 1,463/362,631.

To further characterize our IL-18 PGS model, we once again used the IBD Plexus SPARC cohort. To test the relationship between the IL-18 PGS and IL-18 gene expression levels, we identified 372 UC and CD cases with both genotyping data and gene expression levels measured in gut tissue. Gene expression levels underwent standard batch correction and were transformed into Z-scores. The final model used was: transformed IL-18 expression ∼ IL-18 PGS + Age + Sex + Top 10 Genetic PCs. Analyses were conducted using all CD and UC patients at once (with Disease Status included as an additional covariate for this model), only using UC patients, and only using CD patients (Supplementary Table 3). Additionally, all analyses were conducted only using gene expression levels from colon biopsies at 20cm proximal of the anal verge.

To test the relationship between the IL-18 PGS and clinical variables, we used 6pt Mayo Scores and short-CDAI scores to categorize UC and CD cases respectively into disease activity categories: remission, mild, moderate and severe. We combined these categories to create binary outcomes to estimate PGS relationships in cases with moderate/severe activity against either cases in remission/mild disease activity or remission only. The final binomial model used was: Disease Activity Group ∼ IL-18 PGS + Age + Sex + Top 10 PCs + Years since diagnosis at time of disease activity measurement (Table 4).

We also assessed the relationship between the IL18 PGS with TNF-a treatment failure in a subset of cases. To create a TNF-a treatment failure variable, we used an algorithm to identify CD and UC cases likely to have failed anti-TNF_α_ therapy since few cases had treatment notes detailing actual treatment response. Only cases meeting strict inclusion criteria were evaluated; we excluded cases with unclassified IBD, no EHR data, investigational drug use, less than one year of follow-up, no anti-TNF_α_ therapy, or missing both start and end dates for any biologic treatment. For missing medication dates, we imputed values based on the assumption that biologics are typically administered in a single day and records are updated at each visit. Anti-TNF_α_ treatment failure was assigned to cases that both discontinued anti-TNF_α_ therapy and then switched to another biologic, as the switch suggests non-response or side effects rather than remission or insurance issues. Cases with ongoing anti-TNF_α_ use and no switches were considered not to have treatment failure. Cases with no (or unclear) anti-TNF_α_ use were classified as unknown anti-TNF_α_ therapy failure and excluded from treatment failure analyses. The final binomial model used was: ever TNFa failure ∼ Age + Sex + Years since diagnosis + Years of follow-up + Top 10 PCs

### IL-18 PGS PCA Projection and Correction

In order to calibrate a PGS score on a single individual to be corrected for ancestry and standardized, such that any individual can be compared against a threshold to assign PGS+ status, we employed an approach that trains and applies two linear models. This approach is similar to what has been used previously in the literature^56^. First, we create a principal component space using genome-wide SNP information (∼450k SNPs) from the 1000 Genomes Phase 3 data, which can then be used to derive principal components for any single individual via principal component projection. Next, we train two linear models to predict the mean and variance of the IL-18 PGS model based on only age, sex, and top 10 principal components. To train these two models, we used the UK Biobank data and projected each individual into the 1000G principal component space in order to derive top principal components. To predict the mean of the PGS, the following linear model was used: PGS ∼ Age + Sex + Projected Top 10 PCs. And to predict the variance of the PGS, the following linear model was used along with the predicted mean from the first linear model: (PGS – PGS Predicted Mean)^2 ∼ Age + Sex + Projected Top 10 PCs. The trained linear models, including their coefficients, are shown below (equations I and II).

Next, we used these trained linear models to correct and standardize each individual’s PGS in the UKB and IBD Plexus SPARC datasets. Using the same set of genome-wide markers as was used to generate the 1000G principal component space, individuals from the UKB and SPARC were projected into the 1000G principal component space and principal components were derived. Then, using age, sex, and these projected principal components, the predicted mean and variance of each individual’s IL-18 PGS was determined using the trained linear models below (equations I and II). With these predicted means and variances determined, each individual’s IL-18 PGS was standardized as follows: (Original PGS – Predicted PGS Mean) / sqrt(Predicted PGS Variance). This final, corrected IL-18 PGS was then compared against the determined threshold to assign PGS+ status.

Trained Linear Model Equations:

I. Predicted PGS Mean = 7.013e-07*Age + -1.811e-05*Sex + -8.028e-03*PC1 + -1.973e-02*PC2 + 1.938e-02*PC3 + -9.831e-03*PC4 + -5.032e-04*PC5 + 2.050e-02*PC6 + 3.217e-02*PC7 + 2.018e-03*PC8 + -1.714e-02*PC9 + 3.524e-03*PC10 + 2.255e-03
II. Predicted PGS Variance = 2.524e-04*Age + 6.986e-03*Sex + 3.405e+00*PC1 + 3.226e+00*PC2 + -1.137e+00*PC3 + 3.793e+00*PC4 + - 2.931e-01*PC5 + -1.648e+00*PC6 + -2.608e-01*PC7 + 1.857e+00*PC8 + - 1.329e+00*PC9 + -2.686e+00*PC10 - 1.021e+01

### IL-18 PGS Thresholding Calibration

To determine a threshold to designate IL-18 PGS high/low status, a grid search approach was taken using both IL-18 protein information from IBD Plexus SPARC and IBD risk information from UKB. First, the threshold which leads to the strongest binary PGS predictor of IL-18 Olink levels was determined using SPARC. Specifically, the IL-18 PGS was converted into binary high/low status using a range of cutoffs going from 0.01 to 0.99 percent. Then, to simulate the presence of a real IL-18 high population, we additionally split individuals into binary classifications of either high or low IL-18 using the same cutoff range from 0.01 to 0.99 percent. Each IL-18 PGS binary predictor was then tested against every IL-18 Olink high/low outcome for a total of 9,801 different combinations of tests. The model that was specifically used was the following: IL-18 Olink High/Low ∼ IL-18 PGS High/Low + Age + Sex + Top 10 PCs. Note that the IL-18 NPX values adjusted for IL-18BP were used in this analysis, the IL-18 PGS model corrected for ancestry as described above was used, and the projected PCs from the 1000G setup also described above were used as well. The PGS threshold that lead to the most significant binary IL-18 PGS predictor out of any combination was then chosen as the cutoff to represent IL-18 prediction.

Next, to incorporate IBD risk information, a similar approach was taken but now with the UKB dataset previously described. We once again created high/low IL-18 PGS predictors using cutoffs from 0.01 to 0.99 percent. We then tested these binary IL-18 PGS as predictors of IBD risk with the following model: IBD Risk ∼ IL-18 PGS High/Low + Age + Sex + Top 10 PCs. Once again, the IL-18 PGS model corrected for ancestry was used as well as the projected PCs from the 1000G setup. A total of 99 tests were conducted and the threshold that lead to the strongest IL-18 PGS high/low predictor of IBD risk was then chosen as the cutoff to represent IBD prediction.

Given these two thresholds then, a combined threshold incorporating both was determined by taking the average percent between them. This produced a single IL-18 PGS cutoff that now represented the threshold for determining IL-18 PGS high/low status incorporating both IL-18 protein prediction and IBD risk prediction.

### IL-18 PGS Ancestry Analysis

IL-18 PGS ancestry analyses were conducted in two different ways using the UKB data. First, for analyses looking at the number of individuals that were considered PGS-high given a specific cutoff (Table 5 and Figure S5), the non-UKB-PPP component of the UKB was extracted and then separated into individual ancestry groups. Ancestry groupings were based on self-reported ethnic background (UKB data-field 21000). For analyses using the major ancestry groupings European, African, and Asian, the self-reported individual ethnic groups were combined as follows: British were designated as the European ancestry group, African and Caribbean were designated as the African ancestry group, and Indian, Chinese, and Pakistani were designated as the Asian ancestry group.

Second, for analyses looking at the percent variance of IL-18 NPX (adjusted for IL-18BP NPX) explained by the IL-18 PGS, as well as the per-SNP sensitivity of the IL-18 PGS, the UKB-PPP portion of the UKB dataset was now used (Figures 5, 6, and S6). Specifically, the UKB-PPP dataset used for the discovery GWAS was subset down to three equally sized groups (n=763) of the same major ancestry groupings described above using data-field 21000, European, African, and Asian. Linear regression analyses were then conducted in each equally-sized subset of the UKB-PPP using the following model: IL-18 NPX (adjusted for IL-18BP NPX) ∼ IL-18 PGS. Datapoint 95% confidence intervals were determined empirically using 10,000 bootstraps. The same model and ancestry groups were used for the per-SNP sensitivity analyses. For these analyses, IL-18 PGS models were constructed that incorporated each SNP one at a time ranked by their original GWAS (or conditional GWAS, if appropriate) p-value.

Additionally, for this second set of ancestry analyses, the gnomAD^60^ dataset was also incorporated. Using version 4.1.0, SNPs were looked up in the gnomAD browser (https://gnomad.broadinstitute.org/) and the allele frequencies within each ancestry group were extracted.

## Data Availability

The SPARC IBD data are available upon approved application to Crohn’s & Colitis Foundation IBD Plexus. UKB-PPP individual-level data are available upon approved application to UKB.

## Author Contributions

M.C.T, J.C.M., and E.R.H. contributed to conceptualization. M.C.T, N.R., and C.L.C contributed to data curation and analysis. M.M., J.C.M., and E.R.H. contributed to supervision. M.C.T. and E.R.H. contributed to writing. All authors contributed to revisions.

## Acknowledgements

The authors would like to thank Corneliu Bodea, Zach Fuller, Frédéric Baribaud, David Zammit, Julie Su, Laurence Menard, Gilles Buchwalter, and Aisling O’Hara Hall for their feedback during this work. Research using the UK Biobank Resource has been done under Application Numbers 26041 and 65851.The results published here are in whole or part based on data obtained from the IBD Plexus program of the Crohn’s & Colitis Foundation. The SPARC IBD data are available upon approved application to Crohn’s & Colitis Foundation IBD Plexus (https://www.crohnscolitisfoundation.org/research/plexus).

## References

1. Ramos, G. P. & Papadakis, K. A. Mechanisms of Disease: Inflammatory Bowel Diseases. Mayo Clin. Proc. 94, 155–165 (2019).

2. Chang, J. T. Pathophysiology of inflammatory bowel diseases. N. Engl. J. Med. 383, 2652– 2664 (2020).

3. Verstockt, B., Bressler, B., Martinez-Lozano, H., McGovern, D. & Silverberg, M. S. Time to revisit disease classification in inflammatory bowel disease: Is the current classification of inflammatory bowel disease good enough for optimal clinical management? Gastroenterology 162, 1370–1382 (2022).

4. Landy, E., Carol, H., Ring, A. & Canna, S. Biological and clinical roles of IL-18 in inflammatory diseases. Nat. Rev. Rheumatol. https://doi.org/10.1038/s41584-023-01053-w (2023) doi:10.1038/s41584-023-01053-w.

5. Pizarro, T. T. et al. IL-18, a novel immunoregulatory cytokine, is up-regulated in Crohn’s disease: Expression and localization in intestinal mucosal cells. J. Immunol. 162, 6829– 6835 (1999).

6. Kanai, T. et al. Interleukin 18 is a potent proliferative factor for intestinal mucosal lymphocytes in Crohn’s disease. Gastroenterology 119, 1514–1523 (2000).

7. Leach, S. T. et al. Local and systemic interleukin-18 and interleukin-18-binding protein in children with inflammatory bowel disease. Inflamm. Bowel Dis. 14, 68–74 (2008).

8. de Lange, K. M. et al. Genome-wide association study implicates immune activation of multiple integrin genes in inflammatory bowel disease. Nat. Genet. 49, 256–261 (2017).

9. Kurki, M. I. et al. FinnGen provides genetic insights from a well-phenotyped isolated population. Nature 613, 508–518 (2023).

10. Mokry, L. E. et al. Interleukin-18 as a drug repositioning opportunity for inflammatory bowel disease: A Mendelian randomization study. Sci. Rep. 9, 9386 (2019).

11. Canna, S. W. et al. An activating NLRC4 inflammasome mutation causes autoinflammation with recurrent macrophage activation syndrome. Nat. Genet. 46, 1140–1146 (2014).

12. Canna, S. W. et al. Life-threatening NLRC4-associated hyperinflammation successfully treated with IL-18 inhibition. J. Allergy Clin. Immunol. 139, 1698–1701 (2017).

13. Wada, T. et al. Sustained elevation of serum interleukin-18 and its association with hemophagocytic lymphohistiocytosis in XIAP deficiency. Cytokine 65, 74–78 (2014).

14. Geerlinks, A. V., Dvorak, A. M. & XIAP Deficiency Treatment Consortium. A Case of XIAP Deficiency Successfully Managed with Tadekinig Alfa (rhIL-18BP). J. Clin. Immunol. https://doi.org/10.1007/s10875-022-01236-2 (2022) doi:10.1007/s10875-022-01236-2.

15. Ter Horst, R., et al. Host and environmental factors influencing individual human cytokine responses. Cell 167, 1111–1124.e13 (2016).

16. Schirmer, M., Kumar, V., Netea, M. G. & Xavier, R. J. The causes and consequences of variation in human cytokine production in health. Curr. Opin. Immunol. 54, 50–58 (2018).

17. Liu, C. et al. Cytokines: From clinical significance to quantification. Adv. Sci. (Weinh.) 8, e2004433 (2021).

18. Lin, L. et al. A systems immunology approach to investigate cytokine responses to viruses and bacteria and their association with disease. Sci. Rep. 12, 13463 (2022).

19. Wray, N. R., Goddard, M. E. & Visscher, P. M. Prediction of individual genetic risk to disease from genome-wide association studies. Genome Res. 17, 1520–1528 (2007).

20. Dudbridge, F. Power and predictive accuracy of polygenic risk scores. PLoS Genet. 9, e1003348 (2013).

21. Choi, S. W., Mak, T. S.-H. & O’Reilly, P. F. Tutorial: a guide to performing polygenic risk score analyses. Nat. Protoc. 15, 2759–2772 (2020).

22. Anastasi, F. et al. Proteomic polygenic risk scores of age-related plasma protein levels reveal a role for Metalloproteinase inhibitor 2 (TIMP2) in cognitive performance. Neurobiol. Aging 157, 68–78 (2025).

23. Donoghue, L. J. et al. Integration of biobank-scale genetics and plasma proteomics reveals evidence for causal processes in asthma risk and heterogeneity. Cell Genom. 5, 100840 (2025).

24. Lewis, C. M. & Vassos, E. Polygenic risk scores: from research tools to clinical instruments. Genome Med. 12, 44 (2020).

25. Fahed, A. C., Philippakis, A. A. & Khera, A. V. The potential of polygenic scores to improve cost and efficiency of clinical trials. Nat. Commun. 13, 2922 (2022).

26. Xiang, R. et al. Recent advances in polygenic scores: translation, equitability, methods and FAIR tools. Genome Med 16, 1–14 (2024).

27. Marston, N. A. et al. Endothelial cell-related genetic variants identify LDL cholesterol-sensitive individuals who derive greater benefit from aggressive lipid lowering. Nat. Med. 1–7 (2025).

28. Sands, B. E. et al. Phase 2 trial of anti-TL1A monoclonal antibody tulisokibart for ulcerative colitis. N. Engl. J. Med. 391, 1119–1129 (2024).

29. Sun, B. B. et al. Plasma proteomic associations with genetics and health in the UK Biobank. Nature 622, 329–338 (2023).

30. Bycroft, C. et al. The UK Biobank resource with deep phenotyping and genomic data. Nature 562, 203–209 (2018).

31. Raffals, L. E. et al. The development and initial findings of A study of a prospective adult research cohort with inflammatory bowel disease (SPARC IBD). Inflamm. Bowel Dis. 28, 192–199 (2022).

32. Mbatchou, J. et al. Computationally efficient whole-genome regression for quantitative and binary traits. Nat. Genet. 53, 1097–1103 (2021).

33. Dinarello, C. A., Novick, D., Kim, S. & Kaplanski, G. Interleukin-18 and IL-18 binding protein. Front. Immunol. 4, 289 (2013).

34. Duncan, J. A. & Canna, S. W. The NLRC4 inflammasome. Immunol. Rev. 281, 115–123 (2018).

35. Ghayur, T. et al. Caspase-1 processes IFN-gamma-inducing factor and regulates LPS-induced IFN-gamma production. Nature 386, 619–623 (1997).

36. Devi, S. et al. CARD-only proteins regulate in vivo inflammasome responses and ameliorate gout. Cell Rep. 42, 112265 (2023).

37. Vance, R. E. The NAIP/NLRC4 inflammasomes. Curr. Opin. Immunol. 32, 84–89 (2015).

38. Matico, R. E. et al. Structural basis of the human NAIP/NLRC4 inflammasome assembly and pathogen sensing. Nat. Struct. Mol. Biol. 31, 82–91 (2024).

39. Kayagaki, N. et al. Caspase-11 cleaves gasdermin D for non-canonical inflammasome signalling. Nature 526, 666–671 (2015).

40. Shi, J. et al. Cleavage of GSDMD by inflammatory caspases determines pyroptotic cell death. Nature 526, 660–665 (2015).

41. Chen, G. Y., Liu, M., Wang, F., Bertin, J. & Núñez, G. A functional role for Nlrp6 in intestinal inflammation and tumorigenesis. J. Immunol. 186, 7187–7194 (2011).

42. Elinav, E. et al. NLRP6 inflammasome regulates colonic microbial ecology and risk for colitis. Cell 145, 745–757 (2011).

43. Tuladhar, S. & Kanneganti, T.-D. NLRP12 in innate immunity and inflammation. Mol. Aspects Med. 76, 100887 (2020).

44. Coombs, J. R. et al. NLRP12 interacts with NLRP3 to block the activation of the human NLRP3 inflammasome. Sci. Signal. 17, eabg8145 (2024).

45. Zhang, Z. et al. Protein kinase D at the Golgi controls NLRP3 inflammasome activation. J. Exp. Med. 214, 2671–2693 (2017).

46. Heiser, D. et al. Evaluation of protein kinase D auto-phosphorylation as biomarker for NLRP3 inflammasome activation. PLoS One 16, e0248668 (2021).

47. Zhu, Z. et al. Causal associations between risk factors and common diseases inferred from GWAS summary data. Nat. Commun. 9, 224 (2018).

48. Choi, S. W. & O’Reilly, P. F. PRSice-2: Polygenic Risk Score software for biobank-scale data. Gigascience 8, giz082 (2019).

49. Privé, F., Arbel, J. & Vilhjálmsson, B. J. LDpred2: better, faster, stronger. Bioinformatics 36, 5424–5431 (2021).

50. Mak, T. S. H., Porsch, R. M., Choi, S. W., Zhou, X. & Sham, P. C. Polygenic scores via penalized regression on summary statistics. Genet. Epidemiol. 41, 469–480 (2017).

51. Lloyd-Jones, L. R. et al. Improved polygenic prediction by Bayesian multiple regression on summary statistics. Nat. Commun. 10, 5086 (2019).

52. Ge, T., Chen, C.-Y., Ni, Y., Feng, Y.-C. A. & Smoller, J. W. Polygenic prediction via Bayesian regression and continuous shrinkage priors. Nat. Commun. 10, 1776 (2019).

53. Khera, A. V. et al. Genome-wide polygenic scores for common diseases identify individuals with risk equivalent to monogenic mutations. Nat. Genet. 50, 1219–1224 (2018).

54. Gettler, K. et al. Common and Rare Variant Prediction and Penetrance of IBD in a Large, Multi-ethnic, Health System-based Biobank Cohort. Gastroenterology 160, 1546–1557 (2021).

55. Khera, A. V. et al. Whole-Genome Sequencing to Characterize Monogenic and Polygenic Contributions in Patients Hospitalized With Early-Onset Myocardial Infarction. Circulation 139, 1593–1602 (2019).

56. Lennon, N. J. et al. Selection, optimization and validation of ten chronic disease polygenic risk scores for clinical implementation in diverse US populations. Nat. Med. 30, 480–487 (2024).

57. Martin, A. R. et al. Clinical use of current polygenic risk scores may exacerbate health disparities. Nat. Genet. 51, 584–591 (2019).

58. Wang, Y., Tsuo, K., Kanai, M., Neale, B. M. & Martin, A. R. Challenges and Opportunities for Developing More Generalizable Polygenic Risk Scores. Annu Rev Biomed Data Sci 5, 293–320 (2022).

59. Wang, Y. et al. Global Biobank analyses provide lessons for developing polygenic risk scores across diverse cohorts. Cell Genom. 3, 100241 (2023).

60. Chen, S. et al. A genomic mutational constraint map using variation in 76,156 human genomes. Nature 625, 92–100 (2024).

61. Karczewski, K. J. et al. Pan-UK Biobank GWAS improves discovery, analysis of genetic architecture, and resolution into ancestry-enriched effects. Genetic and Genomic Medicine (2024).

62. Mountjoy, E. et al. An open approach to systematically prioritize causal variants and genes at all published human GWAS trait-associated loci. Nat. Genet. 53, 1527–1533 (2021).

63. Privé, F., Luu, K., Blum, M. G. B., McGrath, J. J. & Vilhjálmsson, B. J. Efficient toolkit implementing best practices for principal component analysis of population genetic data. Bioinformatics 36, 4449–4457 (2020).

64. Kosoy, R. et al. Ancestry informative marker sets for determining continental origin and admixture proportions in common populations in America. Hum. Mutat. 30, 69–78 (2009).

