## Supplementary Figures and Tables for "A proteomic polygenic score to identify IL-18 driven inflammatory bowel disease"

**Supplementary Figures & Tables**

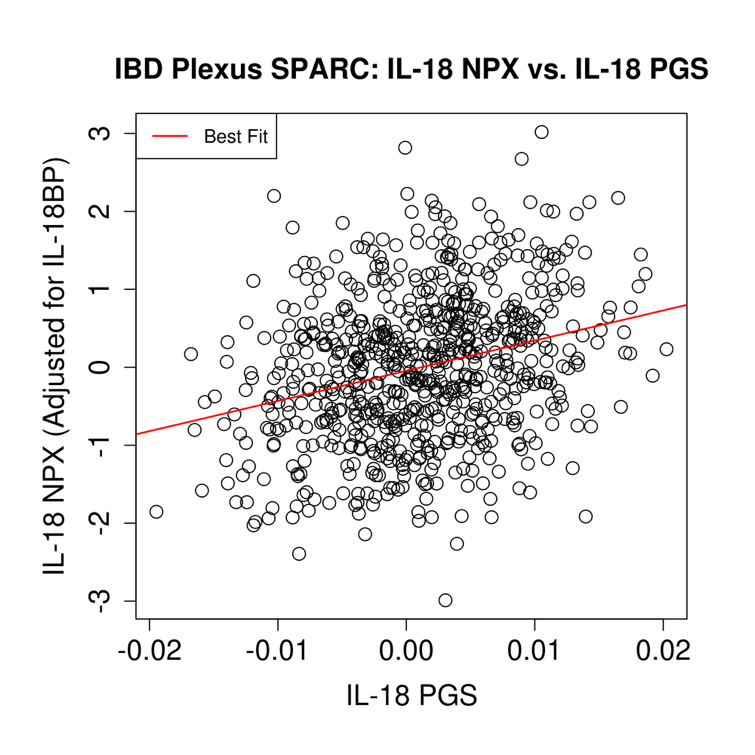

**Figure S1. Prediction of IL-18 using the full conditional mapping model of the IL-18 PGS in SPARC.** A scatterplot showing the relationship between IL-18 NPX values and the full conditional mapping IL-18 PGS model in the IBD Plexus SPARC dataset. IL-18 NPX in the SPARC dataset were adjusted for IL-18BP NPX levels. A total of 731 individuals were analyzed. Beta and *p*-value for the association between IL-18 NPX and the IL-18 PGS were 38.63 and 9.71E-15. The model used for this analysis was: IL-18 NPX ~ IL-18 PGS + Age + Sex + Top 10 PCs. Red line represents the line of best fit for this model.

A)
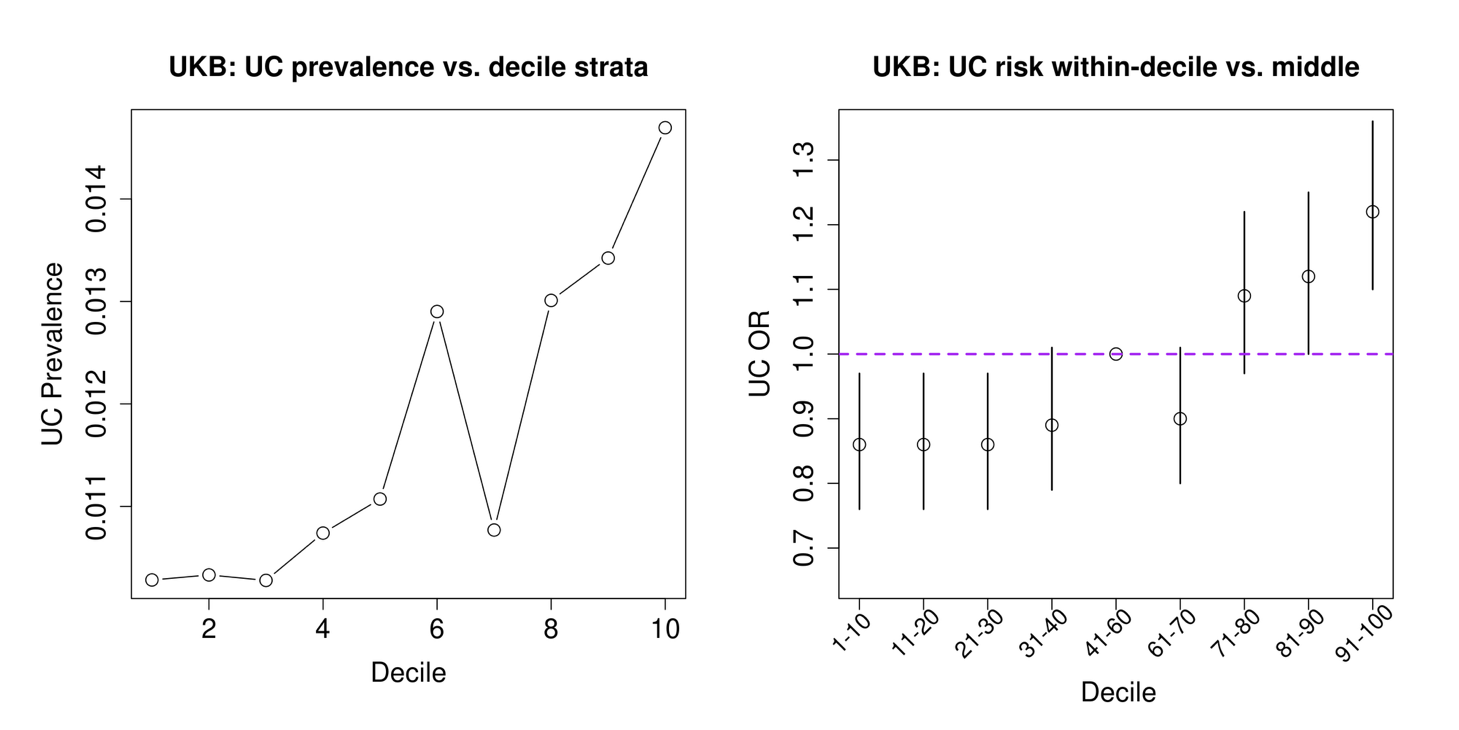

B)
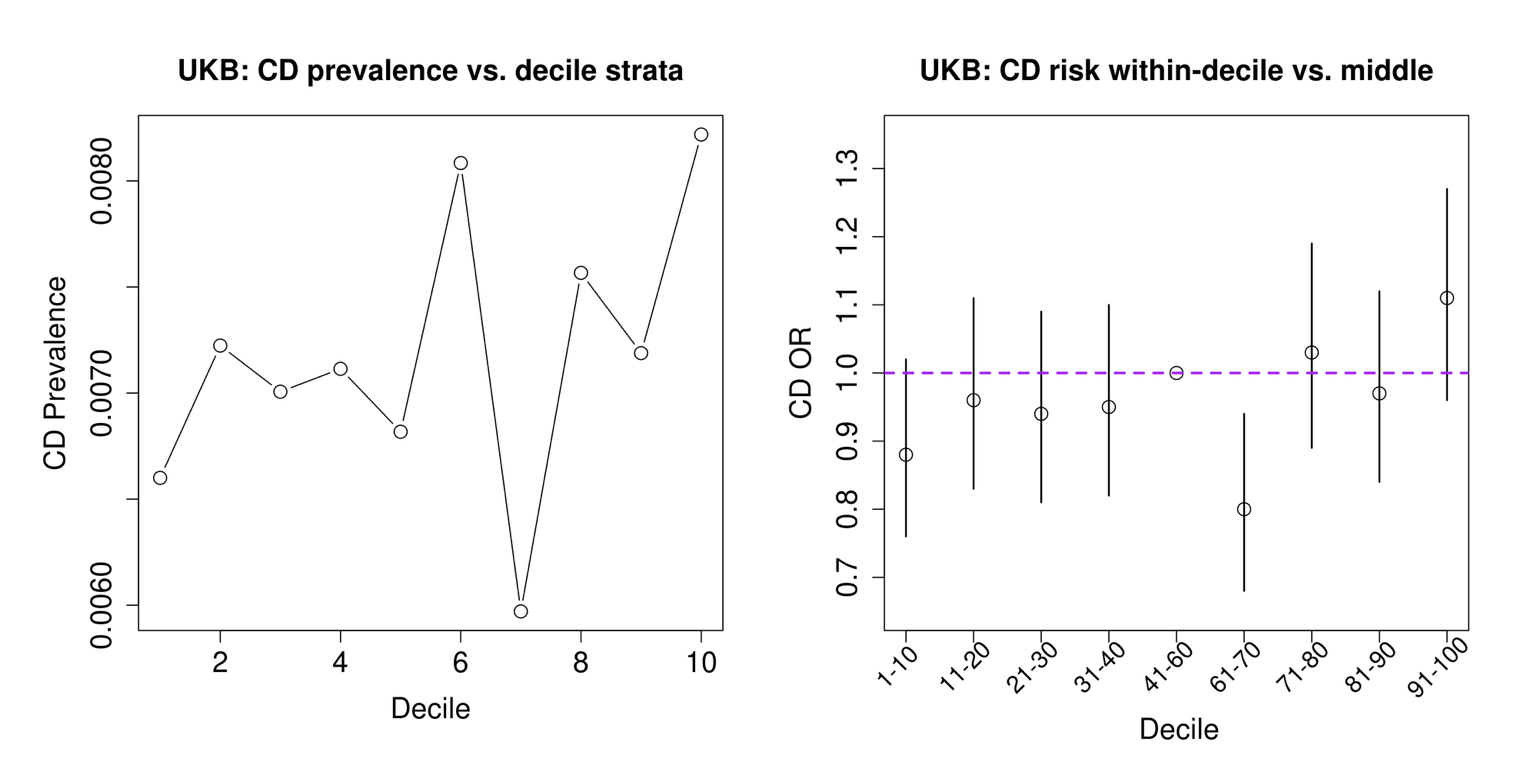

**Figure S2. Prediction of UC and CD using IL-18 PGS Deciles**. Decile plots showing the association between different deciles of the IL-18 PGS and UC (**A**) or CD (**B**) risk when compared to the middle deciles of the dataset. The different deciles of the IL-18 PGS are shown across the *x*-axis. The odds ratio of the association between each decile and UC or CD risk when the middle deciles of 41-60% are treated as the PGS control population are shown on the *y*-axis. Analysis was conducted in the non-UKB-PPP portion of the UKB and UC or CD case status was determined using ICD9/10 codes and self-report (see materials and methods). Error bars represent the 95% confidence intervals. The full model tested was as follows: UC or CD status ~ IL-18 PGS decile status + Age + Sex + Top 10 PCs.

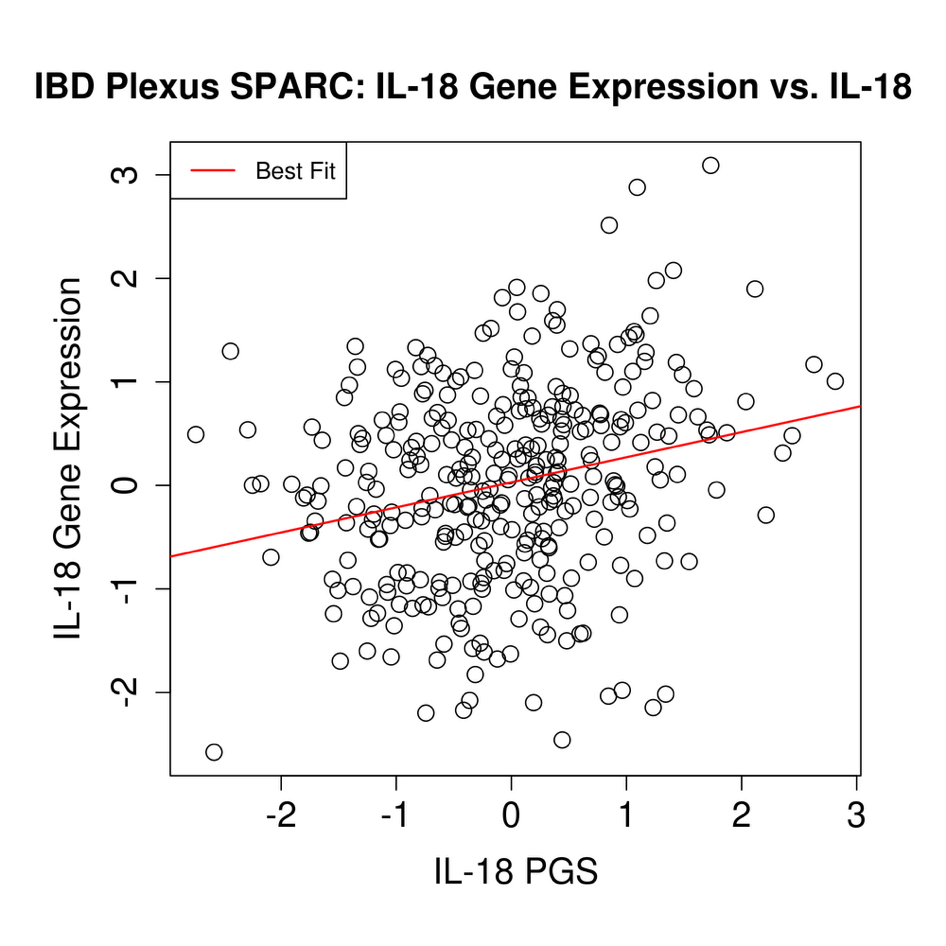

**Figure S3**. **Prediction of IL-18 Gene Expression in Gut Tissue using the IL-18 PGS**. A scatterplot showing the relationship between IL-18 gene expression from gut tissue and the IL-18 PGS in the IBD Plexus SPARC dataset. A total of 327 individuals were analyzed. For individuals with multiple samples, a single random sample was chosen. Gene expression came from colon biopsies at 20cm from the anal verge. IL-18 gene expression underwent batch correction and Z-score transformation. The model used for analysis was: Transformed IL-18 gene expression vs. IL-18 PGS + Age + Sex + Top 10 PCs + Disease Status (UC or CD). Beta and *p*-value for the IL-18 PGS were 0.237 and 1.28E-05. Red line represents the line of best fit for this model. For analysis results solely within UC and CD subsets, see Supplementary Table S2.

A)
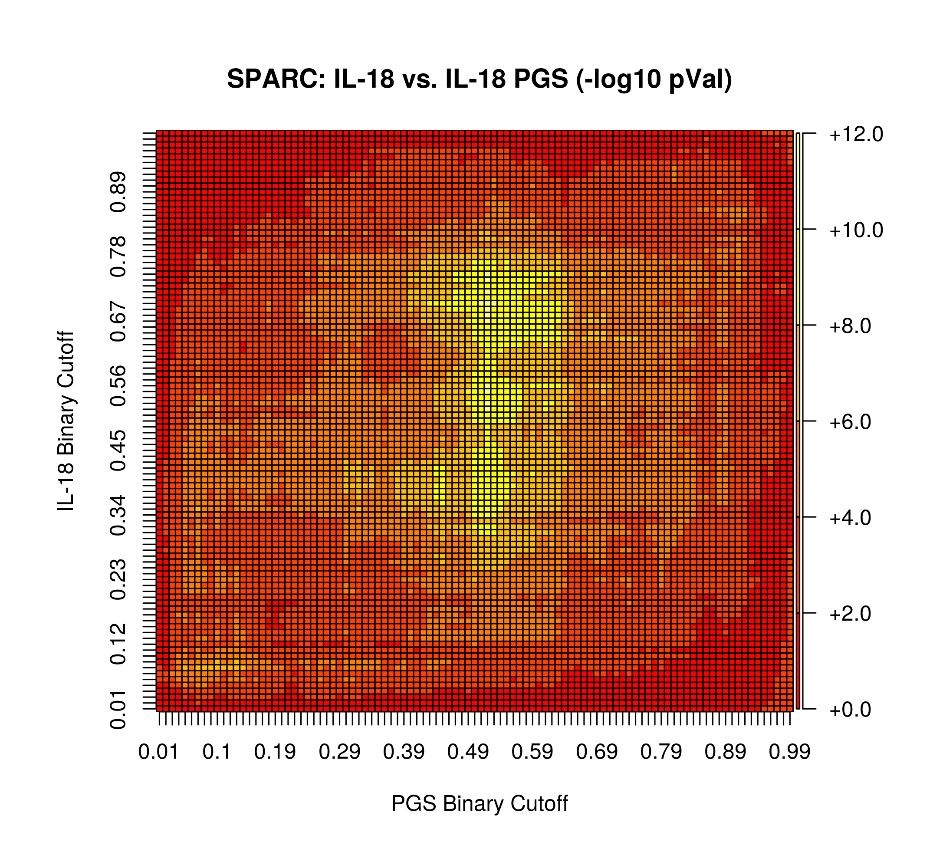

B)
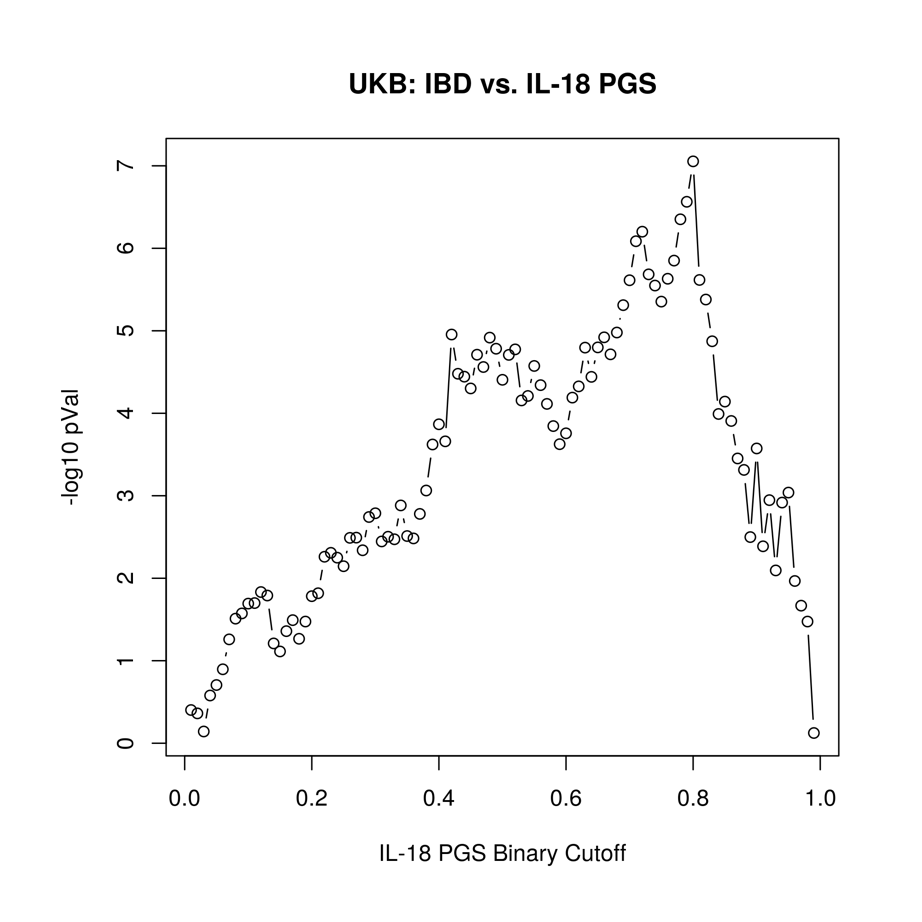

**Figure S4. IL-18 PGS High Threshold Grid-Search Results in IL-18 and IBD. A)** A heatplot showing the association -log10 p-values between binarized versions of the IL-18 PGS and binarized versions of Olink IL-18 (adjusted for IL-18BP) levels. The range of cutoffs used to convert the IL-18 PGS into high/low bins, incremented by 0.01, is shown on the *x*-axis. The range of cutoffs used to convert IL-18 Olink values into high/low bins, incremented by 0.01, is shown on the *y*-axis. The specific model used was as follows: IL-18 Olink High/Low ~ IL-18 PGS high/low + Age + Sex + Top 10 PCs. The IL-18 PGS model used was corrected for ancestry as previously described using 1000G-based projected. Olink IL-18 (adjusted) information came from IBD Plexus SPARC. **B)** A line graph showing the association between binarized versions of the IL-18 PGS and IBD risk across a range of IL-18 PGS high/low cutoffs. The range of cutoffs used to convert the IL-18 PGS into high/low bins, incremented by 0.01, is shown on the *x*-axis. The negative log10 *p*-values of the association between the binarized IL-18 PGS predictors and IBD risk is shown on the *y*-axis. IBD risk information comes from the non-UKB-PPP subset of the UKB dataset.

A)
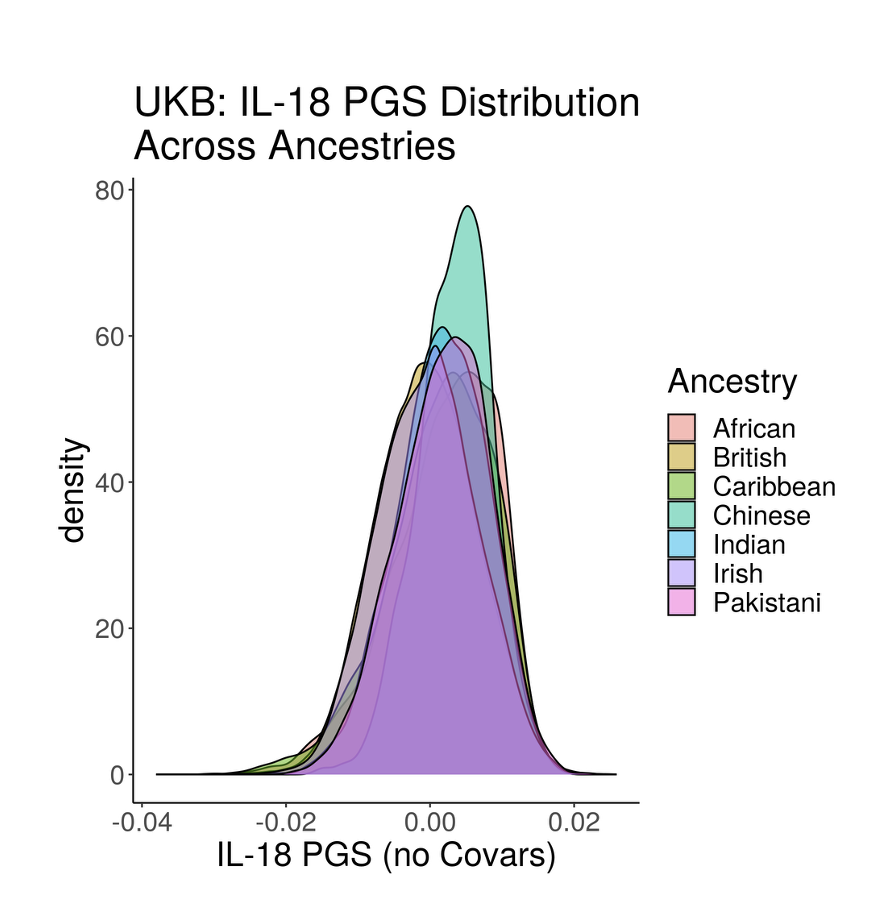

B)
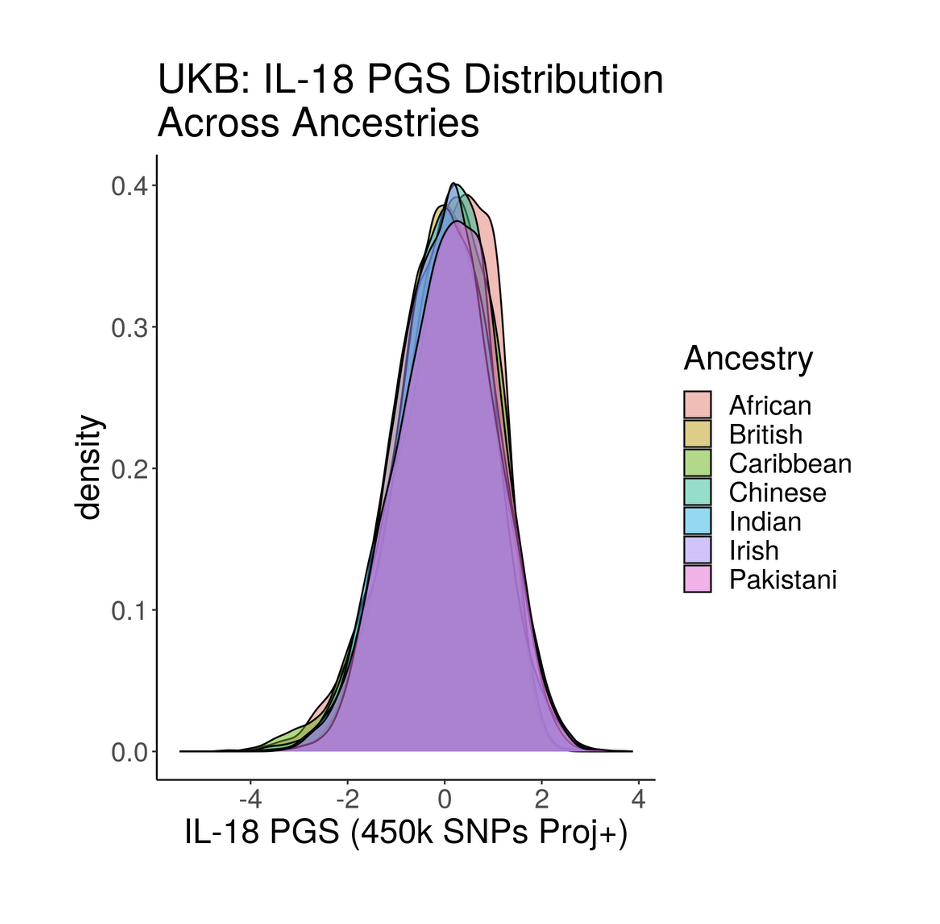

**Figure S5. IL-18 PGS Distributions Across Ancestries and PGS Corrections.** The plots show the IL-18 PGS distribution across UKB ancestry subgroups from two different PGS correction models. The non-UKB-PPP component of the UKB was used. A) shows the distribution of the IL-18 PGS when it has not been corrected by any covariates. B) shows the distribution of the IL-18 PGS when it has been standardized using predicted means and standard deviations from trained linear regression models based on age, sex, and top 10 projected PCs from using 450k SNPs with the 1000 Genomes (see Results and Materials and Methods for further details). Ancestry subgroups were based on UKB self-reported ancestry.

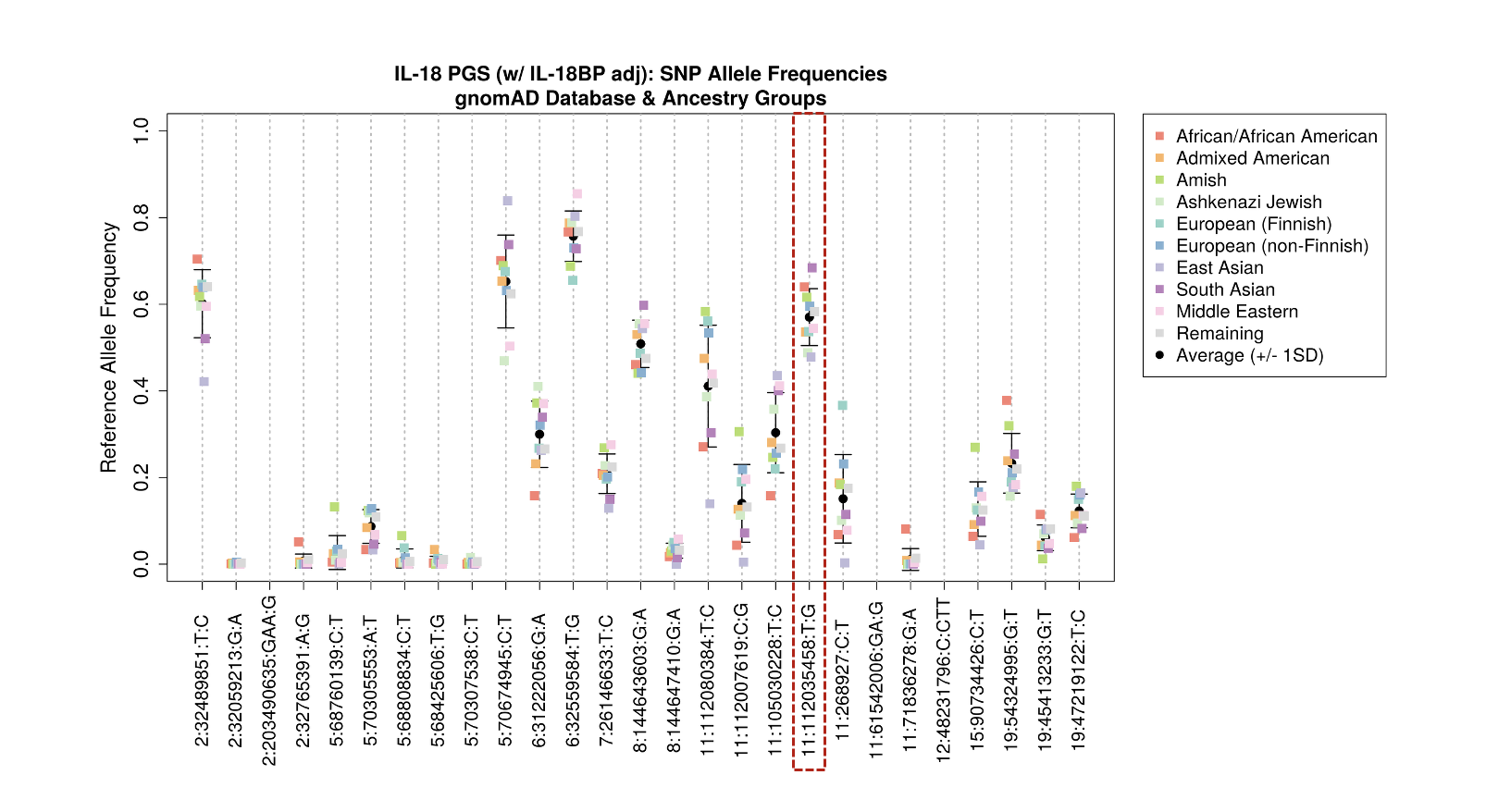

**Figure S6. IL-18 PGS SNP Global Allele Frequencies.** The plot shows global allele frequencies from the gnomAD dataset for each SNP in the IL-18 PGS. The *x*-axis shows each SNP in the IL-18 PGS. The *y*-axis shows the reference allele frequency for each SNP pulled from the gnomAD dataset. Each data point represents a different ancestry group present in the gnomAD dataset. The black dot and error bars represent the average reference allele frequency across every ancestry group, along with one standard deviation, for each SNP. The dotted red rectangle highlights the SNP (rs1946518) where PGS-only r^2^ increases in the full UKB-PPP dataset when it is removed. gnomAD version 4.1.0 was used.

| SNP_ID  (GRCh37) | rsID | Chr | BP (GRCh37) | A1 | A2 | Beta | -log(*p*-Val) |
| --- | --- | --- | --- | --- | --- | --- | --- |
| 11:112080384:T:C | rs10891343 | 11 | 112080384 | T | C | -2.79E-01 | 525.58 |
| 2:32489851:T:C | rs385076 | 2 | 32489851 | T | C | 2.17E-01 | 300.7 |
| 11:112007619:C:G* | rs111311302 | 11 | 112007619 | C | G | -1.75E-01 | 116.67 |
| 11:105030228:T:C | rs1503403 | 11 | 105030228 | T | C | 1.05E-01 | 64.23 |
| 5:68760139:C:T | rs144715385 | 5 | 68760139 | C | T | 2.89E-01 | 62.53 |
| 2:32059213:G:A* | rs191273962 | 2 | 32059213 | G | A | -6.58E-01 | 53.87 |
| 8:144643603:G:A | rs2290414 | 8 | 144643603 | G | A | -8.02E-02 | 46.48 |
| 11:112035458:T:G* | rs1946518 | 11 | 112035458 | T | G | 1.08E-01 | 37.59 |
| 5:70305553:A:T | rs28751879 | 5 | 70305553 | A | T | -1.05E-01 | 33.63 |
| 5:68808834:C:T* | rs149378738 | 5 | 68808834 | C | T | 3.08E-01 | 26.93 |
| 19:54324995:G:T | rs4632248 | 19 | 54324995 | G | T | 7.28E-02 | 26.31 |
| 6:31222056:G:A | rs1793890 | 6 | 31222056 | G | A | -6.17E-02 | 25.16 |
| 11:268927:C:T | rs72882960 | 11 | 268927 | C | T | 6.28E-02 | 21.9 |
| 15:90734426:C:T | rs34560261 | 15 | 90734426 | C | T | -7.01E-02 | 19.31 |
| 5:68425606:T:G* | rs143972019 | 5 | 68425606 | T | G | -2.89E-01 | 18.94 |
| 11:61542006:GA:G | rs111612336 | 11 | 61542006 | GA | G | 5.12E-02 | 18.17 |
| 7:26146633:T:C | rs62446330 | 7 | 26146633 | T | C | -6.01E-02 | 18.12 |
| 11:71836278:G:A | rs2096784 | 11 | 71836278 | G | A | -5.10E-01 | 18.08 |
| 8:144647410:G:A* | rs116635484 | 8 | 144647410 | G | A | 1.16E-01 | 13.4 |
| 12:48231796:C:CTT | rs10675507 | 12 | 48231796 | C | CTT | -4.14E-02 | 12.77 |
| 2:203490635:GAA:G | rs767071613 | 2 | 203490635 | GAA | G | 4.48E-02 | 12.75 |
| 2:32765391:A:G* | rs74883642 | 2 | 32765391 | A | G | -5.64E-01 | 11.32 |
| 6:32559584:T:G | rs9270523 | 6 | 32559584 | T | G | -4.29E-02 | 11.28 |
| 5:70307538:C:T* | rs149371964 | 5 | 70307538 | C | T | 3.19E-01 | 9.42 |
| 19:45413233:G:T | rs1065853 | 19 | 45413233 | G | T | 6.04E-02 | 8.88 |
| 19:47219122:T:C | rs402072 | 19 | 47219122 | T | C | -4.59E-02 | 8.84 |
| 5:70674945:C:T* | rs6453370 | 5 | 70674945 | C | T | 3.51E-02 | 8.47 |

**Table S1. Table of the IL-18 GWAS (adjusted for IL-18BP) lead SNPs.** The table shows detailed information for every SNP in the IL-18 PGS full conditional mapping model based on the IL-18 GWAS that was adjusted for IL-18BP. Summary statistics come from the original IL-18 GWAS. SNPs included due to being conditional lead SNPs are indicated with a star in the SNP_ID column. SNPs are sorted by their -log10 *p*-values. Beta and -log10 *p*-value are from regenie, and beta is in reference to the A2 allele. Human genome build hg37 is used for basepair information.

| SPARC Dataset | N | IL-18 Gene Expression vs. PGS | | |
| --- | --- | --- | --- | --- |
|  |  | Beta | SE | *p*-Value |
| UC+CD | 327 | 0.237 | 0.053 | 1.28E-05 |
| UC | 108 | 0.339 | 0.111 | 3.12E-03 |
| CD | 219 | 0.186 | 0.061 | 2.71E-03 |

**Table S2. IL-18 Gut Tissue Gene Expression vs. IL-18 PGS Models.** The table shows results for analyzing the ability of the IL-18 PGS to predict IL-18 gene expression in gut tissue from different patient subsets of the IBD Plexus SPARC dataset. Three dataset versions were analyzed: one including both UC and CD patients, one using just UC patients, and another using just CD patients. In all three datasets, for individuals with multiple samples, a single random sample was chosen. Gene expression came from colon biopsies at 20cm from the anal verge. Gene expression also underwent batch correction and Z-score transformation. The model used for analysis was: Transformed IL-18 gene expression vs. IL-18 PGS + Age + Sex + Top 10 PCs. For the UC+CD dataset, an additional Disease Status covariate was also included.

| IL-18 PGS Model | SNPs | IL-18 NPX (SPARC) | | IBD Risk (UKB) | |
| --- | --- | --- | --- | --- | --- |
|  |  | Beta | *p*-Value | OR | *p*-Value |
| Original Model | 27 | 0.265 | 9.71E-15 | 1.08 | 2.31E-06 |
| IBD Risk Model | 18 | 0.249 | 9.19E-13 | 1.08 | 4.00E-06 |

**Table S3. Performance comparison of an IBD risk informed PGS model versus the original IL-18 PGS model.** The table shows results from testing an ‘IBD Risk’ version of the IL-18 PGS where SNPs were included only if their IL-18 NPX increasing allele was also the IBD risk increasing allele. This new IL-18 PGS model was tested as a predictor for both IL-18 NPX levels and IBD risk and compared against the original FCM PGS model. IL-18 NPX predict and IBD risk prediction were conducted in the same manner as previously described (Materials and Methods, Table 3, and Figure S1 for further details). For both analyses, PGS were standardized such that the betas for the IL-18 NPX prediction and the odds ratios for the IBD risk prediction could be comparable across PGS models.
